# The role of common and rare genetic variation on adiposity across childhood

**DOI:** 10.1101/2025.05.13.25327505

**Authors:** Katherine A. Kentistou, Jonas Sundfjord, Roya Karimi, Lena R. Kaisinger, Robin J. Hofmeister, Adina E. Lupu, Nicolas Fragoso-Bargas, Yajie Zhao, John A. Tadross, Lukas Steuernagel, Georgina K. C. Dowsett, Sam Lockhart, Jens C. Bruening, Jimmy Liu, Adrian Cortes, Yancy Lo, Jonathan Davitte, Lieven Clement, Alexandra Havdahl, Ole A. Andreassen, Eirik Bratland, Brian Y. H. Lam, Stephen O’Rahilly, Giles S. H. Yeo, Pål R. Njølstad, Zoltán Kutalik, Felix R. Day, Marc Vaudel, John R. B. Perry, Ken K. Ong, Stefan Johansson

**Affiliations:** MRC Epidemiology Unit, Institute of Metabolic Science, University of Cambridge, Cambridge, UK; Mohn Center for Diabetes Precision Medicine, Department of Clinical Science, University of Bergen, Bergen, Norway; Department of Computational Biology, University of Lausanne, Lausanne, Switzerland; MRC Metabolic Diseases Unit, Institute of Metabolic Science – Metabolic Research Laboratories, University of Cambridge, Cambridge, UK; Department of Neuronal Control of Metabolism, Max Planck Institute for Metabolism Research, Cologne, Germany; Wellcome-Wolfson Institute for Experimental Medicine, Queen’s University Belfast, Belfast, UK; Human Genetics and Genomics, GSK, Collegeville PA, USA; Human Genetics and Genomics, GSK, Stevenage, UK; Department of Mathematics, Computer Science and Statistics, Ghent University. Ghent, Belgium; PsychGen Centre for Genetic Epidemiology and Mental Health, Division of Public Health and Prevention,, Norwegian Institute of Public Health, Oslo, Norway; Research Department, Lovisenberg Diaconal Hospital, Oslo, Norway; Promenta Research Centre, Department of Psychology, University of Oslo, Oslo, Norway; Center for Precision Psychiatry, Faculty of Medicine, University of Oslo, Oslo, Norway; Division of Mental Health and Addiction, Oslo University Hospital, Oslo, Norway; Department of Pediatrics and Adolescents, Haukeland University Hospital, Bergen, Norway; University Center for Primary Care and Public Health, Unisanté, University of Lausanne, Switzerland; Computational Biology Unit, Department of Informatics, University of Bergen, Bergen, Norway; Department of Genetics and Bioinformatics, Norwegian Institute of Public Health, Bergen, Norway

**Keywords:** Adiposity, Body mass index, Genetic variant, ALSPAC

## Abstract

Our understanding of the genetic architecture of obesity has primarily been shaped by observations from adult populations with relatively few studies on childhood obesity. To address this gap, we conduct a longitudinal genetic association study in up to ∼600,000 individuals with objectively measured or recalled childhood adiposity-related traits. We identify 624 common variant signals (only 7% are previously reported) associated with childhood adiposity, of which one third have no concordant association with adult BMI. Signals linked to the leptin-melanocortin pathway (*BSX, GNAS*, *LEPR* and *PCSK1*) and incretin-signalling genes (*GIPR* and *GLP1R*) show stronger associations in childhood than in adults, suggesting that childhood provides a more sensitive window for detecting variation in key endocrine and neuropeptide pathways regulating energy balance. This observation is further supported by integrating single-nucleus RNA sequencing data from the human hypothalamus, identifying childhood-specific adiposity-regulating cell populations in the arcuate nucleus and mammillary bodies, indicating distinct neuro-circuits that regulate adiposity only during childhood. Three signals showed parent-of-origin specific associations with childhood BMI, at *KLF14* (maternal-specific), *GNAS* (parental-discordant) and *ZDBF2* (paternal-specific). Finally, we complement these common variant analyses with DNA sequence data in 479,615 individuals, identifying rare protein-coding variation in *ADCY3*, *CALCR*, *MC4R*, *MRAP2*, *POMC* and *MYH13,* all of which demonstrate stronger adiposity associations in childhood than in adults. Collectively, our findings emphasize the value of expanding research on childhood adiposity alongside studies focused on adult obesity measures.

## Introduction

The prevalence of child and adolescent obesity has more than quadrupled over the past 3 decades^1^. These trends are concerning as childhood obesity is linked to many comorbidities^2,3^ and usually tracks into adulthood when it causes long-term adverse health outcomes^4^. Obesity has a pronounced genetic component, with twin and family studies estimating its heritability to be between 40-90%^5^. The largest genome-wide association study (GWAS) for BMI to date identified over 900 genetic loci in over 700,000 adults^6^, and implicated central nervous system (CNS) pathways controlling the hedonic aspects of food intake as a major driver of obesity^7,8^.

Most GWAS loci identified for adult BMI are also associated with obesity in children^9–11^, but there is growing evidence that some genetic factors influence BMI differently between young age and adulthood^12–16^. For example, common variants at *FTO* (rs9939609) and *MC4R* (rs17782313) display biphasic associations with adiposity, which strengthen up to age 20 years and then weaken with age^17^. Other signals, such as rs2767486 in *LEPR*, influence BMI only transiently in infancy^13^. Given these differences, there is an interest in focusing on early life periods to study the genetics of obesity. However, previous GWASs of childhood obesity-related traits have much smaller sample sizes than studies in adults, and most pooled data from across multiple stages of childhood. The largest GWAS on childhood BMI included a discovery sample of 39,620 children aged 2 to 10 years, with targeted replication in up to 22,000 children; it found 25 significant loci^10^. Thus, previous childhood studies have markedly lower power than in adults, and most also lack specificity about the timing of association. Furthermore, they do not address the full allelic spectrum.

To meet these challenges, we apply several new approaches. We perform discovery GWASs for childhood BMI at unprecedented age-granularity and sample size, at 11 timepoints from age 6 weeks to 8 years in a greatly increased analysis of up to 62,276 children from the Norwegian Mother, Father and Child Cohort study (MoBa). We next use a genomic structural equation modelling (gSEM) approach^18^ to combine different childhood adiposity-related traits: MoBa childhood BMI at age 8, UK Biobank recalled relative adiposity at age 10, and a GWAS meta-analysis of age of menarche (a trait closely influenced by early life adiposity^19^). This yielded an effective GWAS sample size of 599,924 individuals. We characterise the age-specific trajectories of association of identified signals on BMI, analyse single cell sequence data to compare their distinct enrichment in specific hypothalamic cell types to adult BMI signals, and use whole genome sequence data to identify child-specific rare variant associations. Our approach uncovers genetic determinants of childhood adiposity at an unprecedented depth.

## Results

### Cross-sectional and aggregation of signals for childhood BMI in MoBa

GWAS was performed in up to 62,276 children from MoBa for age- and sex-standardized BMI (zBMI) at each of 11 timepoints from ages 6 weeks to 8 years (Figure 1a-c, **Supplementary Data 1**). We identified between 7 and 100 conditionally independent genome wide significant (*P*<5×10^-8^) SNPs at each timepoint (total 369 associations) (**Supplementary Data 2**). BMI at age 6 months yielded the most independent SNPs (**Figure 1c**). SNP-based heritability estimates varied with age, mirroring the changes in absolute BMI, increasing after birth to the ‘adiposity peak’ at age 1 year, then decreasing until the age at ‘adiposity rebound’ (**Figure 1c, Supplementary Data 4**). There was only modest genetic correlation between BMI in infancy and childhood (8m-8y: r^2^=0.39) and even lower with adult BMI (8m-Adult: r^2^=0.19). These results illustrate the changing genetic architecture of BMI with developmental stages (Fig 1d).

**Figure. 1.**
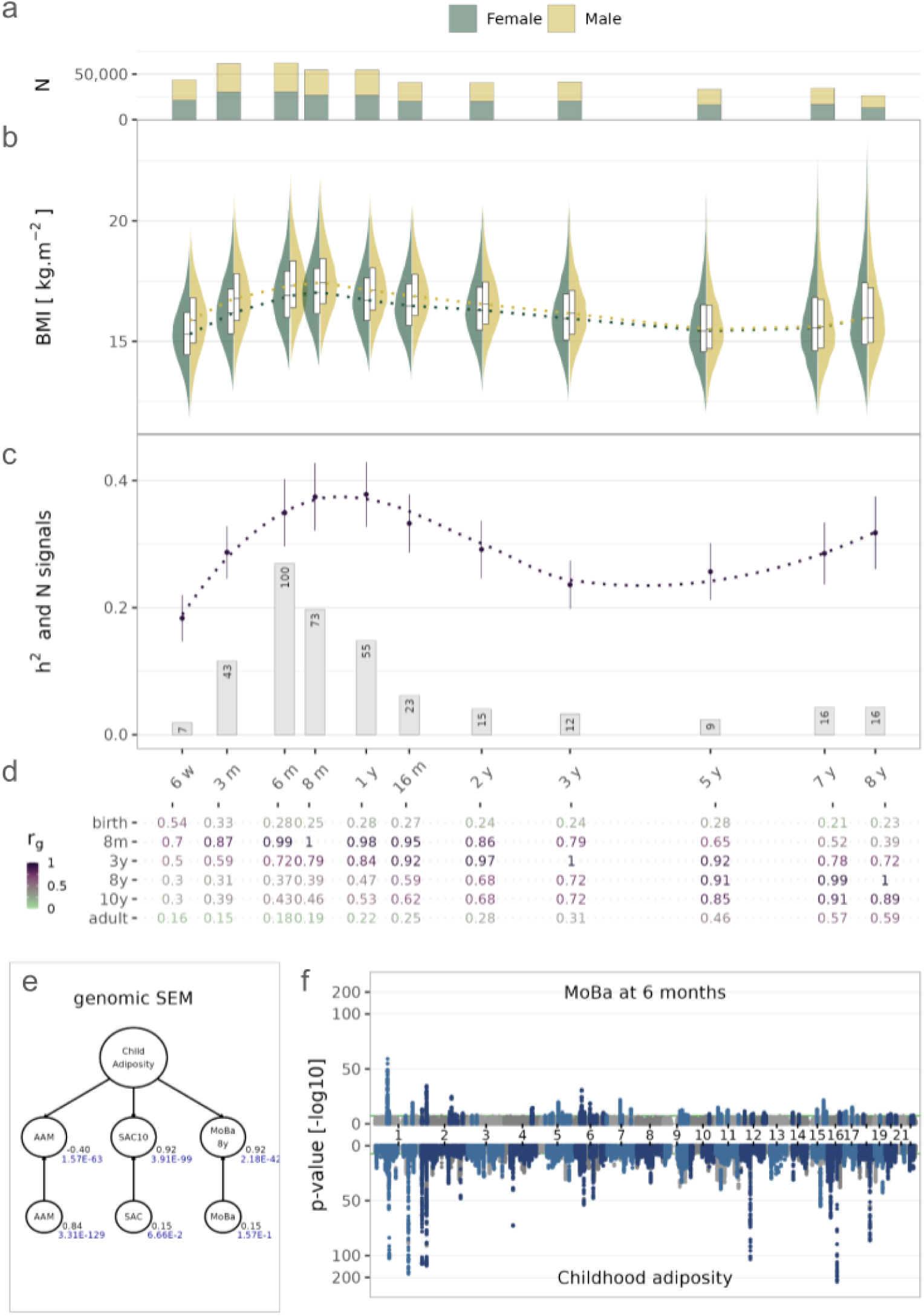
Genetic discovery of variants influencing BMI throughout childhood. (**a**) Number of children included in the GWAS at each timepoint. (**b**) Violin and box plots of the BMI distributions at each timepoint. (**c**) Error bars display SNP-based heritability (h^2^, 95% CI) and its interpolation (dashed line) and columns display the number of independent signals at each timepoint. (**d**) Genetic correlations for BMI at birth, 8 months, 3 years, and 8 years in MoBa, recalled adiposity at 10 (SAC) in UK Biobank, and adult BMI^20^. (**e**) Genomic SEM (gSEM) common factor loadings (higher circles) and residual variance (lower circles) for each contributing factor. (f) Miami plot of the MoBa GWAS at 6 months (upper plot) and the gSEM child adiposity common factor GWAS (lower plot). Genome-wide significant variants are highlighted in blue.

Several of the 369 SNPs identified across the 11 timepoints mapped to overlapping loci, and we aggregated these into 152 independent signals (*R*^2^<0.05); at each signal we retained the most strongly associated SNP at each timepoint (see **Methods**, **Supplementary Data 3**). Of these 152 signals, only 29 (19.1%) were reported in our previous analysis of a subsample of MoBa^13^, and only 12 (7.9%) were reported in the largest GWAS of measured childhood BMI^10^ (**Supplementary Data 3**).

To estimate the variance in BMI explained by common variants, we constructed 2 sets of polygenic scores (PGS) at each timepoint: i) comprising the 152 conditionally independent lead SNPs identified in MoBa (PGS_LeadSNP **Supplementary Data 5**) and ii) a genome-wide score (PGS_GW **Supplementary Data 6**), and compared these to corresponding PGSs for adult BMI^20^ (**Supplementary Data 7**). We then regressed the BMI of children from the independent ALSPAC cohort against our time-stratified PGSs (**Supplementary Data 5-7**). Both the time-stratified PGS_LeadSNP and PGS_GW outperformed their corresponding adult BMI-based PGSs for all timepoints below age 5 years. For example, at 8 months, close to the adiposity peak, the PGS_LeadSNP and PGS_GW explained 7.0% and 10.9% of the variance in BMI in ALSPAC at 8-12 months, respectively, compared to only 1.3% explained by a genome-wide PGS for adult BMI (**Supplementary Figure 1**).

### Using childhood adiposity-related phenotypes to increase discovery

Although the heritability of BMI increased again after age 3 years (**FIgure 1c, Supplementary Data 4**), we identified relatively few signals for BMI at ages 7-8 years due to fewer children at these timepoints in MoBa (**Figure 1b**, **Supplementary Data 2**). To leverage greater power, we constructed a gSEM model by combining data from MoBa childhood BMI at age 8 (MoBa8), recalled comparative adiposity at age 10 (SAC10) in UK Biobank, and a recently published GWAS for age at menarche (AAM)^19^ to estimate a ‘childhood adiposity’ common factor. Almost half of all genetic variants associated with age at menarche (AAM), act through early-life weight-related mechanisms, and there is a moderate inverse genetic correlation between female puberty timing and adult BMI (*R*_g_=-0.35)^21^. The effective sample size of this analysis was 599,924 and it performed well across all quality control and validation metrics (**Supplementary Information**). SAC10 and MoBa8 showed strong factor loadings on the childhood adiposity common factor (both 0.92, **Figure 1e**), while AAM had a moderate negative loading (-0.40, **Figure 1e**), consistent with the genetic correlations between the individual traits.

We performed a GWAS for this childhood adiposity common factor and identified 526 independent (*R*^2^<0.05) genome-wide significant signals (*P*<5×10^-8^), after excluding 117 heterogeneous signals (defined by Q index *P*_Q_<5×10^-8^, **Supplementary Data 8**). Those heterogeneous signals had disproportionately large effects on AAM and relatively weak association with MoBa8 or SAC10 (**Supplementary Figure 2**). Our childhood adiposity common factor showed high genetic correlation with measured childhood BMI in the independent EGG sample^10^ (*R*_g_=0.85). The 526 GWAS signals showed high directional concordance with both measured childhood BMI (472/520, 91%, *P*=3.6×10^-89^) and adult BMI (487/525, 93%, *P*=6.0×10^-101^, **Supplementary Data 8**).

A PGS comprising the 526 signals for the childhood adiposity common factor explained up to 9.2% of childhood BMI (peak age 9) (**Figure 2**) and up to 6.1% of childhood body-fat-percent (peak age 9) in ALSPAC (**Supplementary Data 7**). This performance was higher than similar PGSs based on SAC10 alone or adult BMI on childhood BMI (maximum *R*^2^_SAC10_ =7.5%, *R*^2^_adult_ =7.0%) and childhood body-fat-percent (maximum *R*^2^_SAC10_ =5.5%, *R*^2^_adult_ =4.5%) (**Supplementary Figure 3**).

**Figure. 2.**
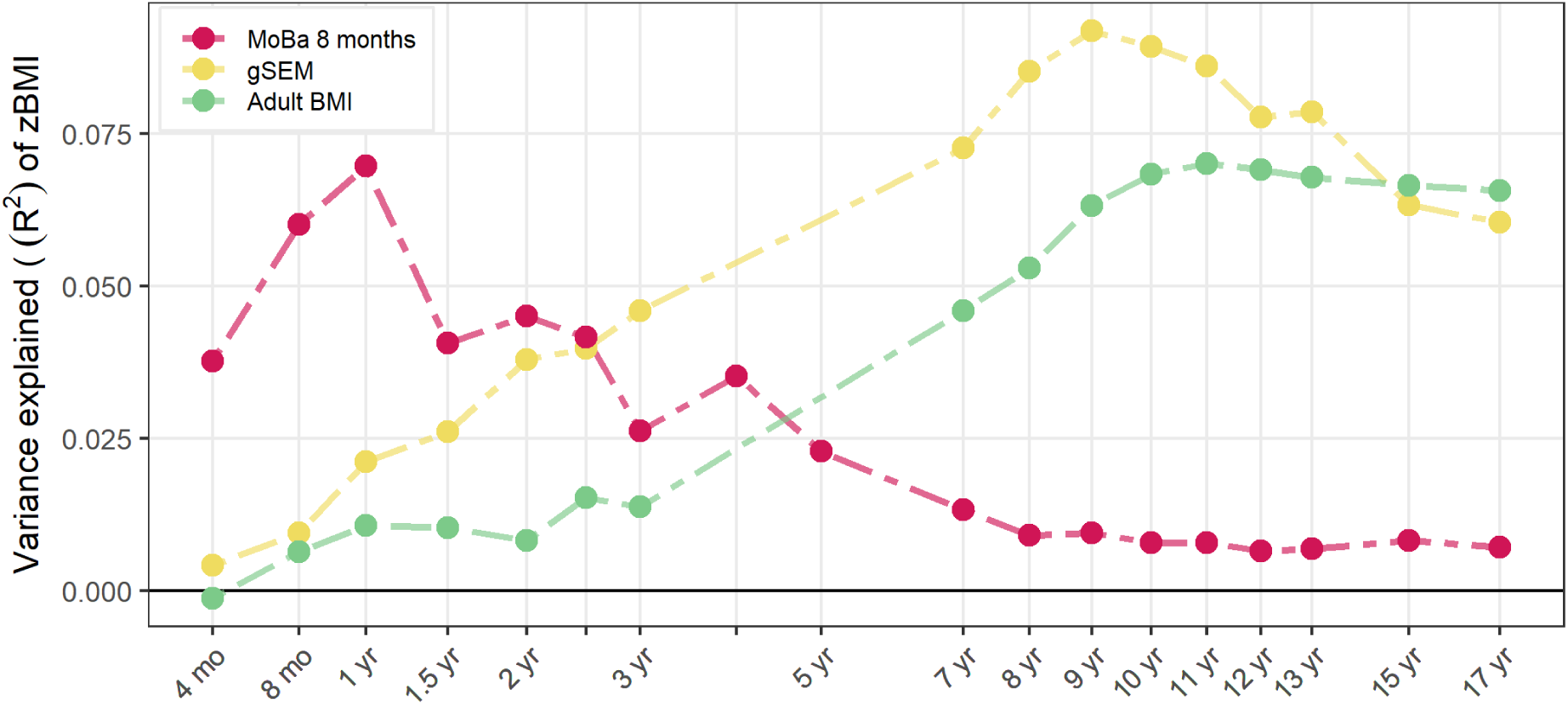
Performance of the childhood adiposity common factor PGS (gSEM) and an infancy-stratified PGS (MoBa 8 months) on BMI across childhood in ALSPAC. Performance is indicated by variance explained of standardized BMI at each age. Performance of an adult BMI PGS^20^ is also plotted for comparison.

### Clustering of variants by their age-related trajectories

To identify age-related clusters of variants for infant and childhood adiposity, we first combined the 152 signals for cross-sectional BMI in MoBa with the 526 signals for the childhood adiposity common factor to identify 624 independent (*R*^2^<0.05) infant or childhood adiposity signals (**Supplementary Data 9**) of which only 7% (n=44) were previously discovered in the two largest studies of childhood BMI^10,16^. We then performed unsupervised clustering of 622 signals (excluding 2 signals with incomplete data) by their age-related trajectories of association with BMI at 12 MoBa timepoints (including at birth), SAC10 from UK Biobank and the childhood adiposity common factor (see **Methods**).

This identified 4 clusters: *Birth-related*, *Transient*, *Early-rise* and *Late-rise* (**Figure 3a), Supplementary Data 10**). These aligned well with the 4 trajectory groups previously reported using a supervised clustering approach in a subset of MoBa^16^. Signals in the Transient cluster (N=115) showed associations with BMI during infancy only, with strongest effects between ages 6 weeks to ∼12 months. Early-rise trajectory signals (N=145) showed associations with BMI from age 6-8 months and persisted into childhood. Late-rise trajectory signals (N=330) showed increasing associations with BMI from age 3 years onwards. The smallest cluster, Birth-related (N=32), showed a “U-shape” trajectory, with associations in the neonatal period and also at age 8-10 years. Examples of selected individual trajectories are shown in **Fig 3b**.

**Figure. 3.**
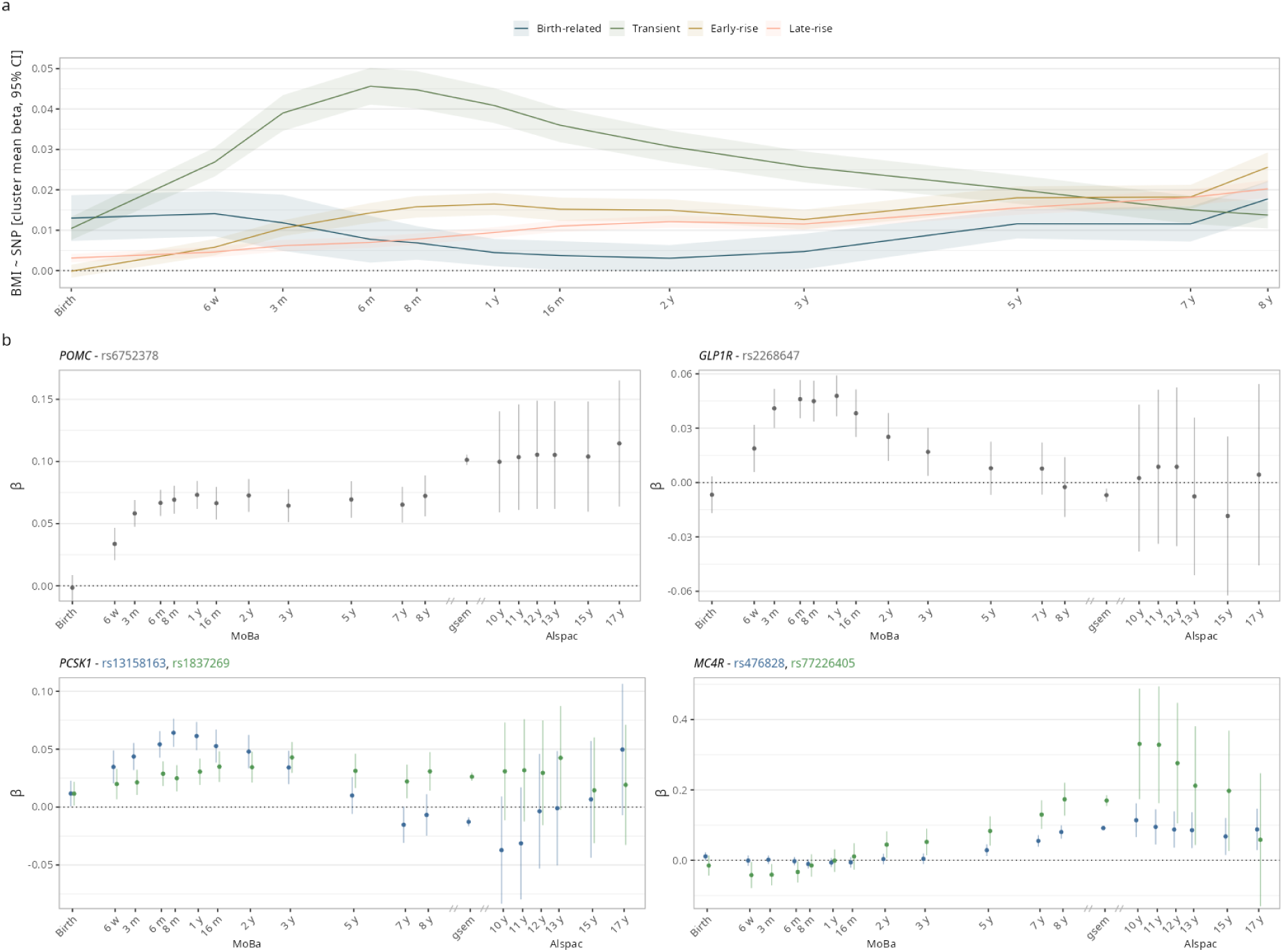
Clustering of variants by their effects on BMI across childhood. (**a**) Mean effect estimates for variants in each trajectory cluster at each timepoint in MoBa. (**b**) Effect estimates (95% CI) on childhood BMI in MoBa and the childhood adiposity common factor (blue lines and shading) and on childhood BMI in ALSPAC (black error bars) for selected individual variants *POMC:* rs6752378-Early rise, *PCSK1*: rs13158163-Transient and rs1837269-Transient, *GLP1R*: rs2268647-Transient, *MC4R*: rs476828-Late rise and rs77226405-Early rise. Further data are in **Supplementary Data 10** and **11**.

We constructed trajectory-specific PGSs and observed confirmatory patterns of association with BMI across childhood in the ALSPAC cohort (**Supplementary Figure 4, Supplementary Data 11**) as well as for individual SNP-trajectories (**Figure 3b**).

### Variant-to-gene mapping implicates childhood adiposity genes

To implicate putatively causal genes at each of the 624 childhood adiposity signals, we applied the ‘GWAS to Genes’ (G2G) analysis framework, which combines data on expression and protein quantitative trait loci (eQTL and pQTL), enhancer maps and common coding variants (see **Methods**)^19^. This approach identified 434 ‘high-confidence’ childhood adiposity genes, defined as the highest-scoring gene at each locus, and supported by concordant evidence from at least 2 independent sources (**Supplementary Data 15**).

Among these high-confidence childhood adiposity genes were several components of the leptin-melanocortin pathway (*BDNF*, *BSX*, *CALCR*, *GNAS*, *LEP, LEPR, MC4R, NTRK2*, *NPY, PCSK1, PCSK2* and *POMC*), which has an established role in appetite regulation and severe early-onset obesity^8^. The signals linked to *BDNF*, *CALCR*, *GNAS*, *LEP, MC4R*, *NTRK2*, *NPY, PCSK2* and *POMC* showed Early-rise and/or Late-rise trajectories of BMI association; *CALCR*, *MC4R* and *PCSK2* harboured multiple signals assigned to both Early-rise and Late-rise trajectories. The signals linked to *BSX*, *LEPR* and *PCSK1* showed Transient BMI associations (**Supplementary Data 16**).

We also implicated incretin pathway components: glucagon-like peptide 1 receptor (*GLP1R*) (1× Transient and 1× unassigned signal), glucose-dependent insulinotropic polypeptide receptor (*GIPR*) (1× Early-rise and 1× Late-rise), and gastrin releasing peptide (*GRP*) (1× Early-rise), which are key regulators of postprandial metabolism and treatment targets for Type 2 diabetes and obesity^22^. Other implicated genes are previously reported obesity susceptibility genes with roles in neuronal development and synaptic function^8^: *CADM1* (Late-rise)*, CADM2* (Late-rise) and *NEGR1* (Early-rise) (**Supplementary Data 16**).

### Putative imprinting effects resolved by trio-analysis

Imprinting is an epigenetic gene silencing process affecting around 1% of human genes resulting in parent-of-origin specific expression, which can be both tissue and development-stage specific^23^. To examine for possible parent-of-origin effects (PoE), we analyzed parental-specific child BMI associations in up to 25,779 unrelated child-mother-father trios in MoBa (see **Methods**, **Supplementary Data 13**). QQ plots showed greater than expected associations among the 18 signals near known imprinted regions, and no inflation among the remaining 606 signals outside imprinted regions (**Figure 4a-b**).

**Figure 4.**
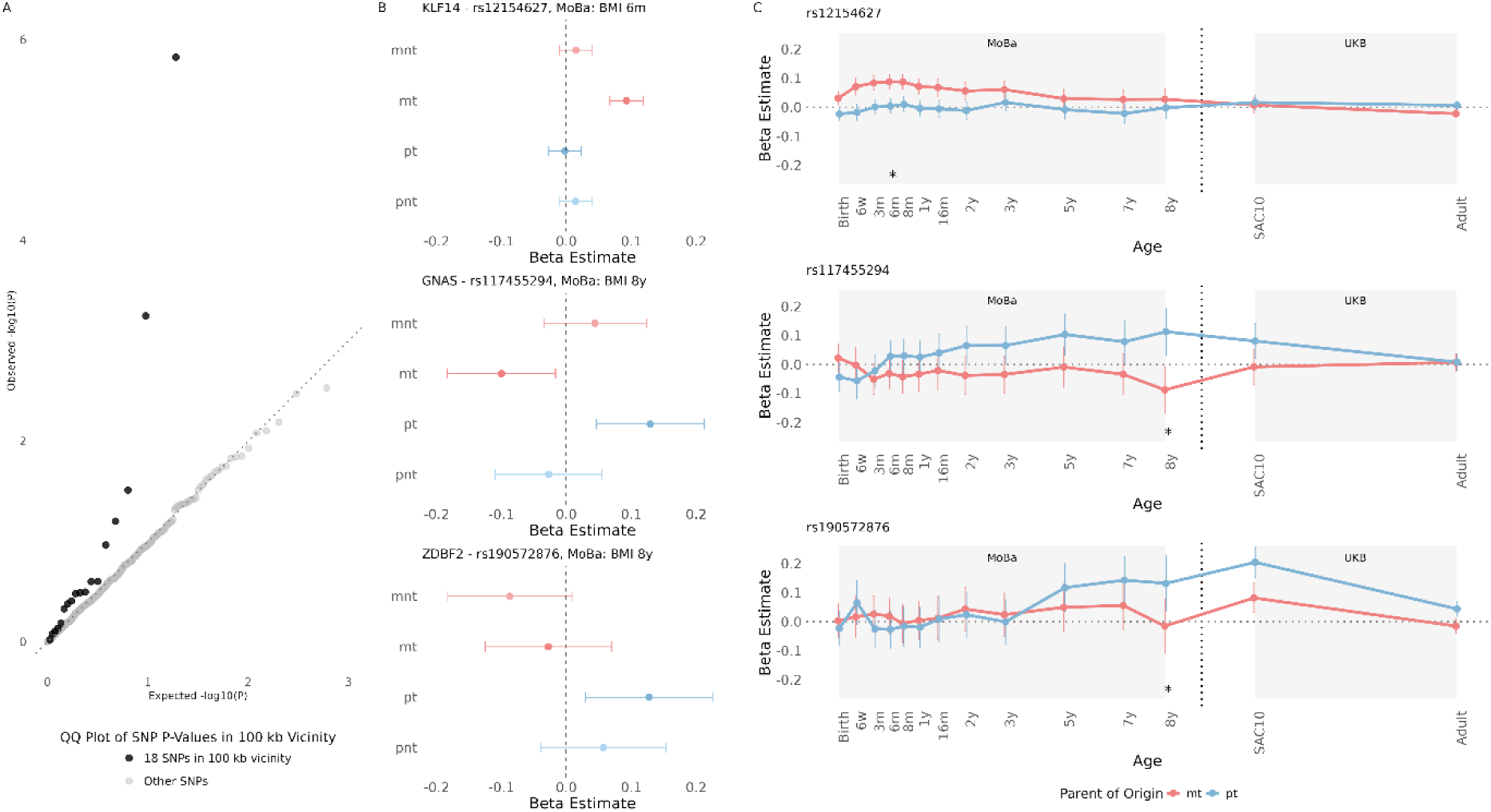
Parent-of-origin effects (PoE) for lead SNPs near known imprinted regions. (**a**) QQ plot showing the distribution of observed versus expected p-values for PoE analyses (the difference between maternal-transmitted (mt) and paternal-transmitted (pt) effects) across all 624 GWAS signals, highlighting inflation among the 18 signals near known imprinted regions but not among the remaining 606 signals. (**b**) Forest plots showing beta estimates and 95% confidence intervals for maternal-transmitted (mt), paternal-transmitted (pt), maternal non-transmitted (mnt), and paternal non-transmitted (pnt) allele effects on BMI at the timepoint with the most significant parent-of-origin effect in MoBa for: rs12154627 (*KLF14*, 6 months), rs117455294 (*GNAS*, 8 years), rs190572876 (*ZDBF2*/*GPR1*, 8 years). (**c**) Trajectories of mt and pt associations by age from birth to adulthood in MoBa and UK Biobank. ‘*’ highlights the timepoint with most significant PoE association. Further data are in **Supplementary Data 13** and **14**.

Evidence for PoE was found at rs12154627-*KLF14* (a Transient signal), on infancy BMI (*P*_MT=_1.3×10^-12^, *P*_PT=_0.75, *P*_diff_=1.5×10^-6^) confirming previous observations in MoBa^16^ and elsewhere^24^. A previously unreported PoE was observed for rs117455294 (Late-rise), located in a differentially methylated region (DMR) at the *GNAS* locus, which showed directionally-discordant effects between maternal-transmitted and paternal-transmitted alleles (BMI at 8 years: *P*_MT=_0.03, *P*_PT=_5.7×10^-3^, *P*_diff_=5.6×10^-4^). A third signal, rs190572876 (Early-rise) showed a novel paternally-transmitted allele association with BMI at 8 years (*P*_MT=_0.75, *P*_PT=_6.4×10^-3^, *P*_diff_=0.03) and was also among the strongest gSEM signals for childhood adiposity (*P*=9.5×10^-49^) located near a conserved imprinted region proximal to *ZDBF2*^25,26^. Paternally-expressed *Zdbf2* is reported to regulate glucose homeostasis and neuropeptide signalling and growth^27^. While G2G prioritised *NDUFS1* at this locus (**Supplementary Data 15**), the paternal-transmitted effects of this signal on childhood BMI and SAC10 make *ZDBF2* a more plausible causal gene.

We replicated and extended these findings by applying a recently described method to infer PoE in large population-based samples without parental genotyping^28^ in up to 109,385 white British UK Biobank participants (**Figure 4c, Supplementary Data 14**). The results further demonstrated the life-trajectories of these PoEs including opposing effects between infancy and adult BMI at the *KLF14* locus, the childhood specific paternal-effect of rs117455294-*GNAS* and the predominantly paternal-effect of rs190572876-*ZDBF2* on both SAC10 and adult BMI.

### Biological pathways enriched for childhood adiposity genes

To identify their common biological pathways, we entered our 434 high-confidence childhood adiposity genes into gene-based pathway enrichment analyses using g:Profiler^29^. This identified 134 enriched biological pathways (**Supplementary Data 17**), of which 5 were specifically enriched for Transient BMI associations, 17 for Early-rise, 24 for Late-rise (a further 3 pathways were enriched for 2 trajectories) (**Supplementary Figure 5**).

As the reference pathways were based on overlapping data, we grouped them using a complete clustering method^19^. This identified 35 enriched pathway groups (**Supplementary Data 18**). Transient BMI signals were enriched in the “*response to growth factor*” pathway group (including *FGFR1*, *SOX11* and *IGF1R*). Early-rise signals were enriched in the “*feeding behaviour*” (including *BSX*, *LEP*, *LEPR*, *MC4R*, *NEGR1* and *NPY*), “*dendrites*”, “*synaptic signalling*” (including *BDNF*, *BRAF*, *NF1* and *NTRK2*), “*neuron projection*” and “*G protein-coupled peptide receptor activity*” (including *CALCR*, *GIPR*, *GLP1R* and *MC4R*) pathway groups. Late-rise signals were enriched in the “*protein kinase activity*”, “*secretion*” (including *GIPR*, *LEP*), and “*transmembrane receptor protein signalling*” (including *FGFR1*, *IGF1R*, *NTRK2*) pathway groups.

### Mendelian Randomisation for metabolic related outcomes

We performed inverse variance weighted Mendelian randomisation (MR) analysis (IVW-MR) to investigate if the different early life BMI trajectories were associated with metabolic and anthropometric outcomes in adulthood (Type 2 diabetes, fat mass, lean mass, body fat percent, and waist-hip ratio (WHR)), with and without correction for adult BMI (**Supplementary Data 12**).

The Transient trajectory showed no association with any outcome *(P*>0.05), suggesting that higher BMI during infancy does not increase the risk for Type 2 diabetes later in life. The other 3 childhood BMI trajectories showed positive associations with most outcomes, which were all attenuated on adjustment for adult BMI. This is consistent with prior MR studies indicating that childhood adiposity associations with later body composition and risk of cardio-metabolic disease are predominantly mediated through their strong association with adult BMI^14^.

### Comparison of effects in childhood and adulthood

Our 624 childhood adiposity GWAS signals included 165 signals previously reported for adult BMI (at *P*<5×10^-8^; including at *FTO*, *MC4R*, *BDNF*, *NEGR1*, *SEC16B* and *TMEM18*), and a further 264 signals with at least nominally significant (*P*<0.05) directionally-concordant associations with adult BMI^20^ (**Supplementary Data 19**). More signals in the Birth-related (84%), Early-rise (79%), and Late-rise trajectories (75%) showed at least nominally significant (*P*<0.05) directionally-concordant associations with adult BMI, than did signals in the Transient trajectory (43%)^14^.

Conversely, 194 of our 624 childhood GWAS signals showed either no association (*P*>0.05, N*=*165) or a nominally significant but directionally-opposite association (N=29) with adult BMI (**Figure 5, Supplementary Data 19**). These signals highlighted genes in the leptin-melanocortin pathway (*BSX, GNAS* and *LEPR*), incretin signalling (*GIPR* and *GLP1R*) and the imprinted loci near *KLF14*. Among a further 10 signals with disproportionately larger effects on childhood than adult BMI were 2 variants at imprinted loci (*ZDBF2* and *DLK1)*.

**Figure 5.**
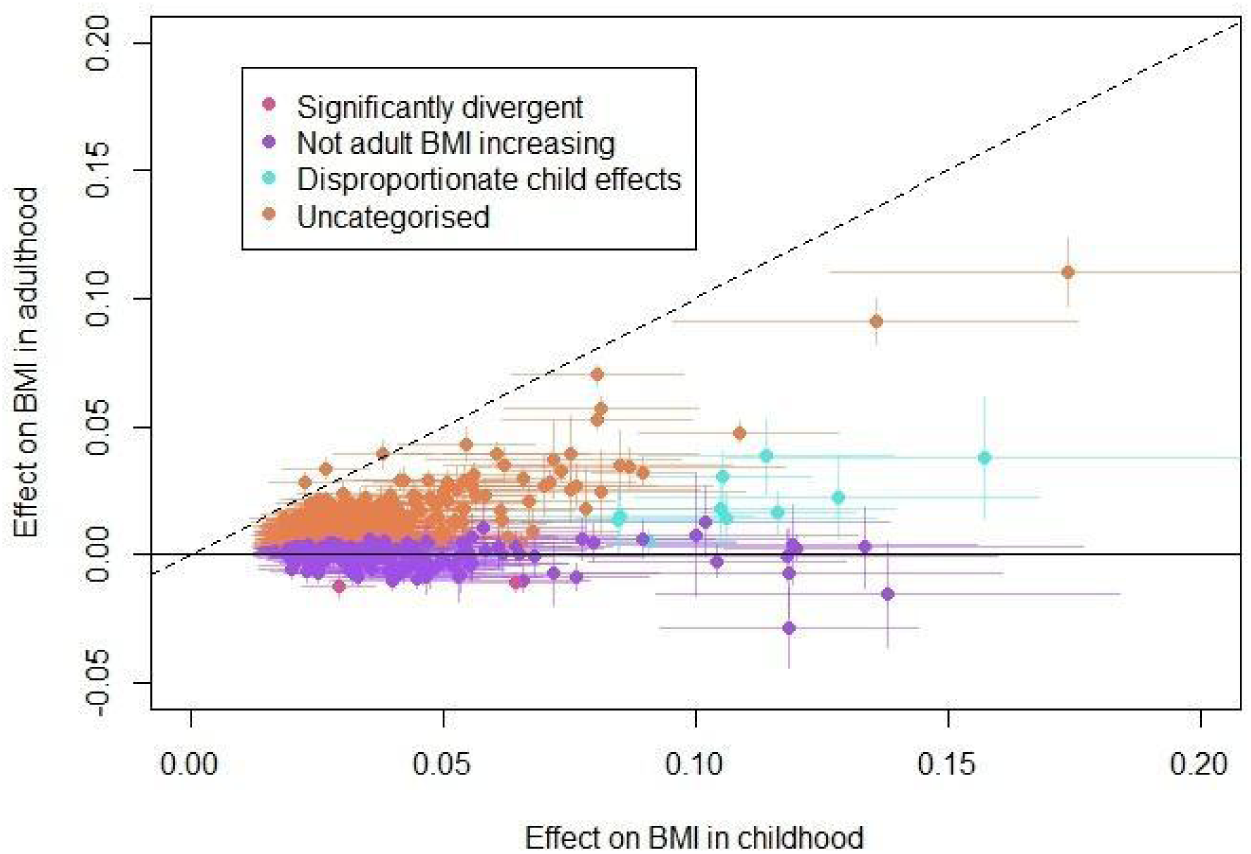
Comparison of childhood and adult BMI effects of identified GWAS signals. Signals identified stratified by their effects on standardised BMI in childhood and in adulthood. Further data are in **Supplementary Data 19**.

Notably, 2 signals showed directionally-opposite, genome-wide significant associations with BMI in children and adults: 1 (of the 2 signals at *PCSK1* (rs13158163) belongs to the Transient trajectory and shows opposing associations with BMI in infancy and adulthood (β_8m_=0.064, *P*_8m_=7.3×10^-25^, β_adult_=-0.011, *P*_adult_=1.3×10^-8^). The roles of common^30^ and rare variants^31^ at *PCSK1* in obesity have been previously reported, however the signal tagged by rs13158163 is restricted to the infancy period and distinct from other common BMI-associated *PCSK1* signals that have persistent association with BMI (rs1837269) (**Figure 3b**). Notably, age-dependent patterns have also been seen in patients with rare biallelic inactivating mutations in *PCSK1*^31^. Such patients show a complex clinical phenotype that includes severe malabsorptive diarrhoea starting in the immediate postnatal period, various endocrine dysfunctions, and later hyperphagic obesity.

### Hypothalamic neuron populations enriched for childhood adiposity genes

To investigate whether distinct neuronal pathways might regulate childhood and adult adiposity, we analysed human hypothalamic single-nucleus RNA sequencing (snRNAseq) data (HYPOMAP)^32^. Our filtered childhood adiposity GWAS signals were enriched in 260 of the 452 reported hypothalamic cell populations (at *P*<0.05/452). All of these 260 were neuronal populations and 19 of them were not also enriched for adult BMI GWAS signals (**Figure 6**, **Supplementary Data 20, Supplementary Figure 6**). Five of these 19 ‘child-specific cell populations’ mapped to the arcuate nucleus (ARC), which plays key roles in energy homeostasis, somatic growth and puberty onset^33^, and a further 5 mapped to the mammillary bodies (MAM) (**Figure 6**).

**Figure 6.**
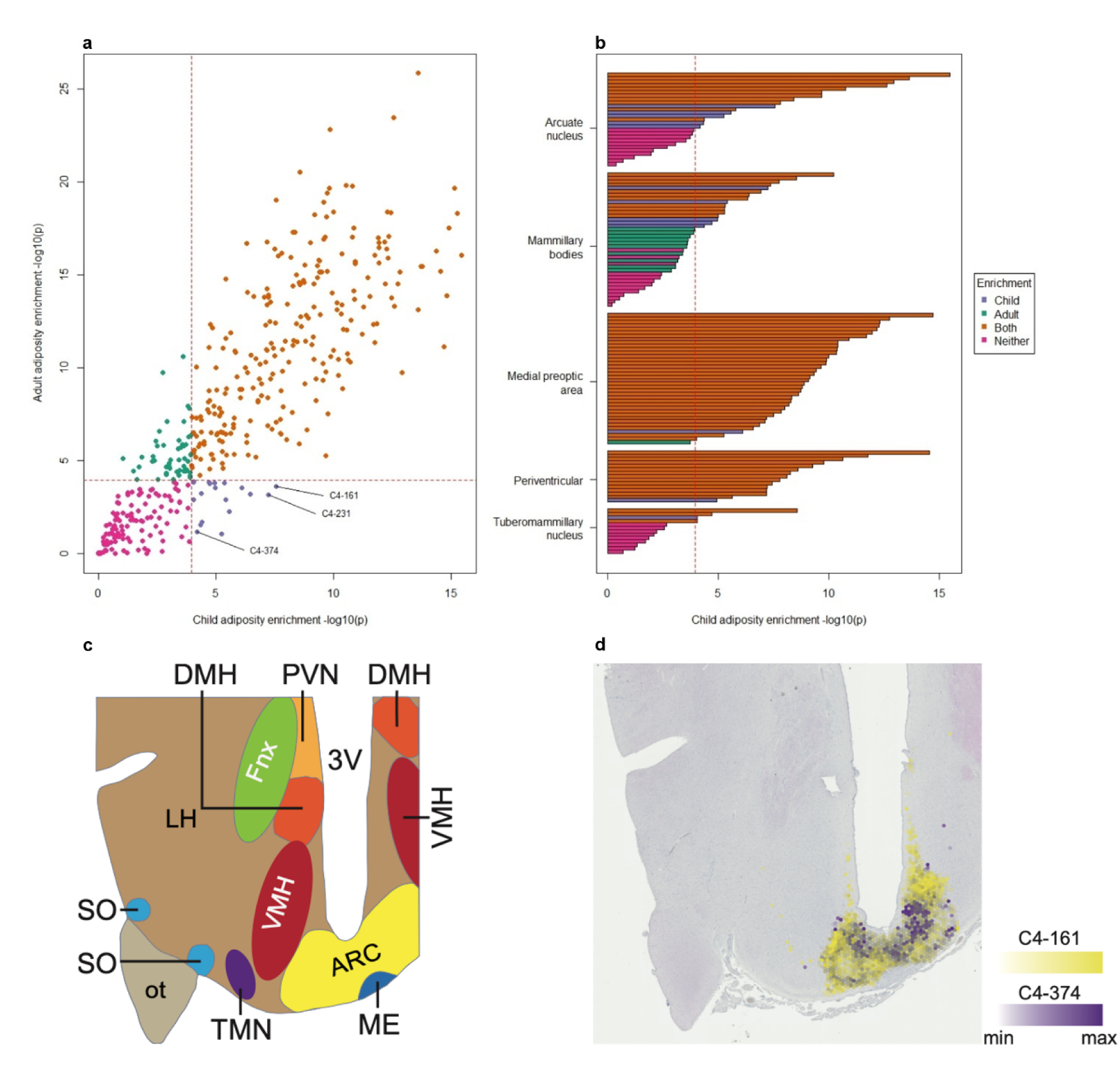
Hypothalamic cell populations specifically enriched for childhood adiposity genes. (**a**) Points represent the 452 HYPOMAP reported hypothalamic cell populations plotted by their enrichment for adult BMI (y-axis) and childhood adiposity associations (x-axis). Blue points are indicating the 19 cell populations enriched only for child adiposity. (**b**) Brain regions containing the child-specific cell populations (blue columns); other cell populations in these regions are indicated by other colours. Dashed lines indicate the Bonferroni-corrected threshold (*P*<0.05/452). (c) Regional annotation of the human hypothalamus (ARC – arcuate nucleus; DMH – dorsal medial hypothalamus; VMH – ventral medial hypothalamus; PVN – paraventricular nucleus; TMN – tuberomammillary nucleus; LH – Lateral hypothalamus; SO – supraotic nucleus; Fnx – fornix; ME – median eminence; 3V – 3^rd^ ventricle; ot – optic tract) (d) Estimated spatial localisation of C4-161 (Mid-1 GABA-6 IL13RA1 GHRH) and C4-374 neurons (Mid-2 GABA-GLU-3 POMC CALCR) within the human hypothalamic arcuate nucleus. Relevant data are in **Supplementary Data 20 and 21**.

Of our high-confidence childhood adiposity genes (identified through G2G), 9 were informative marker genes that contributed strongly to cell-type classification of the 5 child-specific ARC cell populations (see **Methods**). These included components of the leptin-melanocortin (*CALCR*, *LEPR* and *POMC*) and incretin pathways (*GLP1R* and *GRP*), and the imprinted gene, *DLK1* (**Supplementary Data 21**). Similarly, 11 childhood adiposity genes were informative marker genes for the 5 child-specific MAM cell populations. These also included components of the leptin-melanocortin (*BDNF*, *CALCR*, *MC4R* and *PCSK1*) and incretin pathways (*GIPR* and *GRP*).

The highest ARC cell population enriched for childhood adiposity GWAS signals was C4-161 (Mid-1 GABA-6 IL13RA1 GHRH), which is marked by expression of the childhood adiposity genes, *LEPR*, *GLP1R* and *DLK1*. The highest MAM cell population enriched for childhood adiposity GWAS signals was C4-231 (Post-1 GLU-1 EBF2 PLEKHH2), which is marked by expression of *BDNF* and *GIPR* (**Supplementary Data 21**).

### Whole-genome sequencing identifies rare variants affecting childhood adiposity

We analysed whole-genome sequence data in up to 479,615 all-ancestry UK Biobank participants. We performed gene burden tests by collapsing rare variants (MAF<0.1%) within 2 non-overlapping predicted functional categories: (1) high-confidence protein truncating variants (PTVs) and (2) predicted deleterious missense variants with a REVEL^34^ score ≥0.7.

Six genes were associated with SAC10 at a Bonferroni-corrected threshold (*P*<2.9×10^-6^, 0.05/17,265 tests; **Figure 7**, **Supplementary Data 22, Supplementary Figure 7**). These included *POMC*, *CALCR* and *MC4R*, which confirmed our previous report in a smaller UK Biobank sample with whole exome sequencing^35^. Additionally, in *ADCY3*, both rare heterozygous deleterious missense variants (beta=0.177, *P*=4.4×10^−10^, N=575) and PTVs (beta=0.319, *P*=6.4×10^−10^, N=173) conferred larger SAC10. Homozygous LOF of *ADCY3* was previously linked to severe obesity in adults^36,37^. Furthermore, rare heterozygous deleterious missense variants in *MRAP2* conferred larger SAC10 (beta=0.29, *P*=2.2×10^−6^, N=123), consistent with previous findings in hyperphagic obese children and adults^38^. Notably, *ADCY3*, *CALCR*, *MC4R*, *MRAP2* and *POMC* encode central components of the leptin-melanocortin pathway. Finally, we observed a novel association with SAC10 at *MYH13* (beta =-0.1, *P*=2.5×10^−6^, N=1,237 PTV carriers), which encodes a myosin heavy chain that reportedly localises to extraocular and laryngeal muscles^39^.

**Figure 7.**
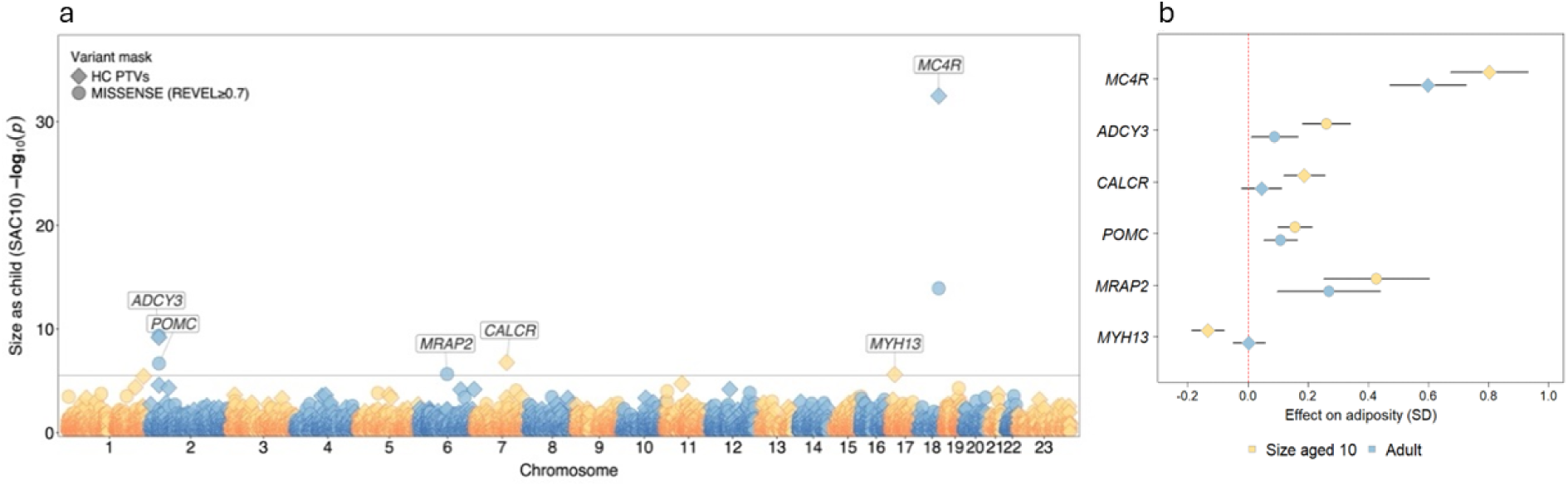
Rare variant gene burden associations with childhood adiposity (SAC10). **a**) Manhattan plot showing genome-wide gene associations. Genes passing the multiple-test corrected threshold (*P*<2.9×10^-6^) are labelled. Predicted variant function masks are indicated by point shapes. b) Identified gene burden associations with childhood adiposity (SAC10) and their associations with adult BMI. Relevant data are in **Supplementary Data 22** and **23**.

Notably, PTVs in *MYH13* showed no association with adult BMI (*P*>0.05). Indeed, although the units of effects were not directly comparable, stronger rare variant associations with SAC10 than adult BMI were apparent for all 6 genes, for example at *ADCY3*: missense variants (*P*_SAC10_=6.4×10^−10^ vs *P*_adultBMI_=0.027); PTVs (*P*_SAC10_=4.4×10^−10^ vs *P*_adultBMI_=7.6×10^−4^) and *MRAP* missense variants (*P*_SAC10_=2.2×10^−6^ vs *P*_adultBMI_=2.2×10^−3^) (**Figure 7, Supplementary Data 22, Supplementary Figure 7**). This observation persisted when we extended association testing at these genes to additional variant masks and other obesity-related traits in UK Biobank (**Supplementary Data 23**).

## Discussion

We report by far the largest GWAS for child adiposity-related traits to date. In addition to novel data on the largest single sample with childhood BMI (from MoBa), we leveraged UK Biobank data on recalled childhood adiposity and on puberty timing to generate an effective sample size of 599,924 individuals for childhood adiposity. This enabled the discovery of 624 independent signals for childhood adiposity, far more than previously reported (25 signals in 61,111 children in EGG^10^, and 46 signals in a subset of MoBa^16^) and comparable to findings for adult BMI (941 signals in >700K individuals)^20^.

Notably, nearly a third (196/624, 31%) of our GWAS signals for childhood adiposity had no concordant effect on adult BMI (*P*>0.05 or directionally discordant). This finding is supported by our demonstration of childhood-specific adiposity-regulating cell populations in the hypothalamic ARC and MAM, suggesting a possible link between recollective memory and energy homeostasis. In both tissues, these childhood-specific cell populations were marked by expression of components of the leptin-melanocortin and incretin pathways. Conversely, we recently reported that of 843 GWAS signals associated with adult BMI in UK Biobank, 114 signals (13.5%) appeared to have ‘adult-specific’ effects^40^. Notably, we did not identify any ARC cell population with adult-specific effects on adiposity.

Our high-confidence childhood adiposity genes included 12 leptin-melanocortin pathway genes, incretin pathway genes, and other obesity susceptibility genes supported by reported evidence from animal models^8^. Two of the highlighted leptin-melanocortin pathway genes were highlighted only by childhood-specific signals (*BSX* and *GNAS*). Others contained multiple independent signals with differing effects on adult BMI. For example, of the 4 independent signals for childhood adiposity linked to *LEPR*, only 1 had a concordant effect on adult BMI. Similarly for incretin receptor genes, of our 2 signals linked to *GIPR*, only 1 appeared to alter adult BMI and none of the *GLP1R* signals was associated with BMI beyond early childhood.

Similarly, in our WGS analysis, it is notable that only 1 of our 6 rare variant-associated genes (*MC4R*) was also identified in a similar analysis of adult BMI in the UK Biobank, which identified 8 other genes^41^, including novel determinants of adult-onset obesity. Five of the 6 genes we highlighted using WGS encode central components of the leptin-melanocortin pathway and have been described or implicated as rare monogenic causes of severe obesity^8^. Here, we provide robust population-based evidence for rare heterozygous variant effects (which have not been previously reported for *ADCY3* and *MRAP2*) and note stronger associations for all genes with SAC10 than with adult BMI.

Our findings also highlight the value of integrating evidence from multiple child obesity-related traits to produce robust findings with high concordance on independent validation. Although we excluded heterogenous gSEM GWAS signals, which appeared primarily driven by puberty timing, some remaining child adiposity signals were linked with high confidence to established regulators of sex hormone secretion: *GNRH1* (rs7832399, Late-rise) encoding a key hypothalamic molecule in the hypothalamic-pituitary-gonadal axis, and *FGFR1* (rs881301, Early-rise) in which mutations cause hypogonadotropic hypogonadism. Reassuringly, these 2 gSEM signals were not driven by confounding due to primary effects on puberty timing as they showed weak associations with AAM (*P*=3.5×10^−2^, *P*=5.1×10^−4^, respectively) and showed concordant associations with childhood BMI at earlier pre-pubertal timepoints (*P*_12m_=7.9×10^−5^, *P*_7y_=7.7×10^−3^, respectively). We note a further Transient adiposity signal was linked to the sex hormone receptor signalling gene *ESR1* (rs1294093; *P*_8m_=5.4×10^−9^). These findings raise the intriguing prospect that the temporary physiological activation of sex hormone secretion during infancy (‘mini-puberty’) might have downstream effects on adiposity^42,43^.

We highlight 2 unique aspects of our study design. First, the multiple repeated BMI measurements in MoBa allowed us to discriminate with fine resolution between different childhood age effects. We found that time-stratified PGS performance peaked in relatively narrow age-windows, illustrating the value of choosing age-matched signals and weights in future studies aiming to predict childhood adiposity or explore its causal relationships.

Second, the availability of child-mother-father trios in MoBa provided unique insights into the role of genomic imprinting on childhood adiposity. It was originally proposed that imprinting evolved to allow the expression of conflicting parental-specific interests on offspring fetal growth (growth-restrictive maternal interests, and growth-promoting paternal interests)^44^. This concept was recently extended to include parental conflict over puberty timing, which impacts the child’s dependence on maternal resources^45^. Our findings further extend this model to childhood adiposity. The growth and metabolic effects of *GNAS* and *KLF14* are well described in humans, and our findings provide new insights into the complex life-long consequences of PoE at these loci. We demonstrate that the PoE at the *GNAS* locus seems confined to childhood, and the PoE trajectory at the maternally-expressed *KLF14* locus shows opposing effects on BMI during infancy and adulthood. While paternally-expressed *Zdbf2* has been reported to regulate early feeding and growth with transient effects in mice^27^, our study is the first to suggest a paternal-only transient association at this region in humans. The imprinted *GPR1*-*GPR1AS*-*ZDBF2* region is well-conserved between human and mouse genomes and further studies are needed to clarify the role of *ZDBF2* in humans. The idea of life-course dependent regulation is also supported by our finding of genes with multiple adiposity signals, which have differing associations at different timepoints, such as *LEPR* and *GIPR*.

We acknowledge some limitations of our approach. While the inclusion of data on puberty timing increased statistical power, larger studies with measured BMI during early childhood are needed to confirm their primary effects on adiposity. We included data on childhood adiposity only from northern European ancestry cohorts. Despite increasing population diversity in available genomics data, this is mostly restricted to studies of adults. Finally, our data on human hypothalamic single-nuclei (HYPOMAP) derived from autopsy material from donors aged 60+ years^32^. Future samples from childhood donors or age-specific animal models would inform whether apparent childhood-specific effects are driven by changes in gene expression or other processes.

In conclusion, these findings emphasize the value of expanding research on childhood adiposity alongside studies in adults. Large-scale genetic studies of childhood adiposity provide richer evidence for energy homeostasis mechanisms compared to studies of adult BMI, suggesting that childhood may provide a more sensitive window for detecting variation in key endocrine and neuropeptide pathways regulating energy balance.

## Methods

### MoBa GWAS

The Norwegian Mother, Father and Child Cohort Study (MoBa) is a pregnancy-based ongoing cohort study consisting of approximately 114,500 children, 95,200 mothers and 75,000 fathers, enrolled from 50 hospitals across Norway from 1999 to 2008. In 41% of the pregnancies invited, the pregnant women consented to participation. The establishment of MoBa and initial data collection was based on a license from the Norwegian Data Protection Agency and approval from The Regional Committees for Medical and Health Research Ethics. The MoBa cohort is currently regulated by the Norwegian Health Registry Act. The current study was approved by The Regional Committees for Medical and Health Research Ethics.

Children’s height and weight were measured by trained nurses during routine checkups at 6 weeks, at 3, 6 and 8 months, and at 1, 1.5, 2, 3, 5, 7 and 8 years of age. These values as reported by parents in questionnaires were processed to compute standardized BMI values at the different timepoints as described in Helgeland *et al.*^16^. Briefly, outliers were removed from growth curves, missing values preceded and followed by at least 2 measurement points were imputed, and BMI values were standardized by sex taking gestational duration as covariate using the generalized additive model for location, scale and shape (GAMLSS) v5.1-7 (<u>gamlss.com</u>).

Genotyping data were acquired collectively by multiple Norwegian research groups on various genotyping platforms and are managed by the Norwegian Institute of Public Health (NIPH). Information on the different genotyping batches is available from the NIPH (github.com/folkehelseinstituttet/mobagen), this study was conducted on the version 1.5 of the genotypes. Quality control of the genotypes was conducted by the PsychGen team, details on the pipeline are available in the respective publication^46^ and code repository (github.com/psychgen/MoBaPsychGen-QC-pipeline). GWAS was conducted at each timepoint using Regenie v. 3.2^47^ as available from Bioconda^48^ and operated using SnakeMake^49^.

### Genomic SEM common factor models

Genomic SEM (gSEM) is a 2-step structural equation modelling approach. First, the empirical genetic covariance matrix and its sampling covariance matrix of included traits are estimated using an extended version of LD-score regression^50^, which takes GWAS summary statistics as input. The regression accounts for potential sample overlap. Second, the user specifies a structural equation model that can omit or include the effects of individual SNPs. Parameters are then estimated by minimising the discrepancy between the model-implied genetic covariance matrix and the empirical covariance matrix obtained in the first step. These analyses were conducted using the gSEM R package^18^ (R version 4.0.2, gSEM version 0.0.5).

First, for each input GWAS, we performed quality control, only including SNPs with minor allele frequency (MAF) >0.1% and imputation information (INFO) score >0.5. As the gSEM software runs in R, which can represent numbers as non-zero only if >=5×10^-324^, we reset the *P*-values of SNPs which were below that value, to this value. Summary statistics for each trait were reformatted using the “munge” function. Second, we ran multivariable LD-Score regression using the “ldsc” function using LD scores and weights for populations of European genetic ancestry^50^. Third, we used the estimated genetic covariance matrix and its associated sampling covariance matrix from the step above to fit a common factor model using Diagonally Weighted Least Squares (DWLS) estimation by applying the “commonfactor” function. We used DWLS estimation as it is recommended over maximum likelihood estimation by the developers of gSEM.

We aimed to predict a single common “latent” factor, i.e. childhood adiposity, and used the following 3 GWASs as inputs: 1) Data on recalled age at menarche (AAM) was taken from the European meta-analysis as described in Kentistou *et al*^19^. Briefly, up to 632,955 European genetic-ancestry female participants were imputed to at least 1000 Genomes reference panel density, and genetic variants were tested for associations with AAM in linear regression models, including age and study-specific covariates. 2) MoBa childhood BMI at age 8 years (MoBa 8, N=26,418), as described above. 3) Data on recalled comparative adiposity (‘size as child, age 10’; SAC10) were available in up to 444,345 White European genetic-ancestry participants from the UK Biobank^35^. The trait (field 1687) is based on the question, “When you were 10 years old, compared to average, would you describe yourself as:” with the following possible answers: “thinner”, “plumper”, “about average”, “do not know”, and “prefer not to answer”. Participants who responded “do not know” or “prefer not to say” were excluded unless, at a later instance, the changed their answer to “thinner”, “plumper” or “about average”. The trait was recoded and analysed as a continuous variable (0=thinner, 1=average, 2=plumper). Association testing was conducted using BOLT LMM^51^.

We tested how well this model fits the assumption of a single “childhood adiposity” common factor using the Comparative Fit Index (CFI) and standardised root mean square residual (SRMR); these are the only 2 indexes computed by the genomic SEM software when the minimum of 3 traits are included. The CFI indicates the extent to which the proposed model fits better than a model that allows all phenotypes to be heritable but genetically uncorrelated. CFI values range from 0 to 1, with values >0.9 considered an acceptable model fit. SRMR is calculated as the standardised root mean squared difference between the model-implied and observed correlations in the model-implied and empirical covariance matrixes. Low SRMR values <0.1 indicate small discrepancy and acceptable model fit^52^. In instances when a model presented with a Heywood case, where the standardised factor loading for an input trait exceeds 1, and the indicator has a negative residual variance, we, instead of using the “commonfactor” function, used a user-specific function imposing a model constraint to keep the residual variance above 0, as advised by the developers of gSEM^18^.

We then performed a GWAS to identify genetic variants associated with the common factor generated by each model. To do so, we first standardised the summary statistics for each trait included in each model using the “sumstats” function, combining the rescaled and aligned data for each trait into a single, listwise deleted file. We treated all traits as continuous and retained insertion-deletion variants (“keep.indel=TRUE”). Next, we used the “commonfactorGWAS” function to generate summary statistics for each SNPs effect on the common factor. We used DWLS estimation as it is recommended over maximum likelihood estimation by the developers of gSEM^18^.

We estimated the expected sample size for the common factor GWAS using the equation:

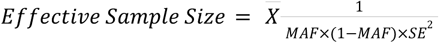

Where 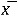 denotes the mean, *MAF* is the minor allele frequency, and *SE* is the standard error of the effect estimate^53^. We restricted MAFs to between 10% and 40% to produce stable sample size estimates as recommended by the developers of gSEM.

Exclusion of heterogenous GWAS signals: Some identified common factor signals may have disproportionately large effects on AAM and have relatively weak association with childhood BMI or SAC10. For each SNP, genomic SEM generates a Q index p-value (pQ), indicating heterogeneity in its associations with the contributing individual traits. Almost all of the signals exhibiting significant heterogeneity (pQ<5×10^-8^) showed highly significant association with AAM but not with any individual childhood adiposity trait (**Supplementary Figure 2**). The only exception was rs663129 (in Models 2 and 3), which showed genome-wide significant associations with SAC10, MOBA7 and MOBA8, but only nominal (and directionally inconsistent) association with AAM. As rs663129 maps to *MC4R*, a well-known obesity gene, and showed strong associations with all adiposity phenotypes, we retained this signal. Further details on gSEM are described in the **Supplementary Information**.

We used LDSC^50^ to calculate the genetic correlation (Rg) between the “childhood adiposity” common factor and childhood BMI from an independent cohort. Data on childhood BMI, measured in children between 2 and 10 years of age, were available in up to 39,620 European genetic-ancestry participants from the EGG consortium^10^.

### Genetic analyses in ALSPAC

Polygenic risk scores (PGS) were calculated for participants in the Avon Longitudinal Study of Parents and Children (ALSPAC)^54,55^. The study initially invited pregnant women resident in Avon, UK with expected dates of delivery between 1st of April 1991 and 31st of December 1992. A total of 14,541 pregnancies were enrolled, resulting in 14,676 foetuses, of which 13,988 children were alive at 1 year of age. An additional 906 pregnancies were enrolled when the children were 7 years old, bringing the total sample size for analyses using data at 7 years and onwards to 15,477 pregnancies. These pregnancies resulted in 15,658 foetuses, with 14,901 children alive at 1 year of age. Initially, 14,203 unique mothers were enrolled in the study, with an additional 630 women recruited when the children were 7 years old, totalling 14,833 women. Partners of the mothers were also invited to participate, with 12,113 partners having been in contact with the study, and 3,807 partners currently enrolled.

We calculated PGS for 7,873 unrelated children with genotyping data from the ALSPAC cohort. Heights and weights between 4 months and 5 years of age were measured in the study clinic in a 10% subsample called the Children in Focus (CiF). For measurements from 7 years of age, the whole cohort was invited to attend the study clinic. BMI measurements were standardized against the British 1990 growth reference for body mass index^56^, Fat percentage was calculated as total body fat (measured by dual-energy X-ray absorptiometry) divided by total body weight. The study website contains details of all available data through a data dictionary and variable search tool (www.bristol.ac.uk/alspac/researchers/our-data).

PGS comprising conditionally independent lead SNPs (PGS_LeadSNP) were calculated for each child by summing effect sizes estimates multiplied by the number of alleles and dividing by the number of variants considered. For time-specific PGSs in MoBa, a set of 152 SNPs was used, corresponding to conditionally independent lead SNPs across all timepoints, and the PGS of a given timepoint was computed using the effect size estimates of these 152 SNPs in the GWAS of MoBa BMI at that timepoint. If a variant was not present in the ALSPAC dataset, a variant in high LD (r^2^ > 0.6) was used as proxy if available.

Genome-wide polygenic scores (PGS_GW) were constructed using LDpred2-auto^57^. SNPs used in the score calculation were limited to those available in the recommended reference panel (HapMap3+), the ALSPAC dataset, and the base GWAS. Individual risk scores were computed for children in ALSPAC from the PGS generated by LDpred2-auto using plink2^58^. Associations between individual variants and childhood BMI in ALSPAC were estimated as described for the MoBa-cohort above.

### Signal identification and GWAS gene prioritisation

To identify independent GWAS signals and prioritise causal genes at the resulting loci, stemming from the timepoint-specific MoBa and gSEM GWAS, we used the “GWAS to Genes” (G2G) pipeline as described in Kentistou et al.^19^ and summarised below.

GWAS summary statistics were filtered to retain variants with a MAF>0.1%. Quasi-independent genome-wide significant signals were initially selected on the basis of distance, within ±1Mb. Secondary signals within these loci were further selected using conditional analyses (GCTA; version 1.93.2)^59^. This was done using an LD reference derived from a random subsample of 25,000 participants of the UKB study. Primary signals were then supplemented with unlinked (*R*^2^<5%) secondary signals, whose association statistics did not overtly change in the conditional models and signals were mapped to proximal NCBI RefSeq genes, within ±500kb.

Independent signals and closely linked SNPs (*R*^2^>0.8) within the associated loci were annotated if they were coding variants within the identified genes or if they mapped within known enhancers of the identified genes, via activity-by-contact (ABC) enhancer maps^60^. Signals were also annotated with their physically closest gene. Gene-level associations were then determined via Multi-marker Analysis of GenoMic Annotation (MAGMA^61^) by collapsing all coding variants within a gene. Colocalisation analyses between the GWAS and expression- or protein-quantitative trait loci (eQTL or pQTL) data were also performed using the SMR-HEIDI (summary data–based Mendelian randomization-heterogeneity in dependent instruments) method (version 0.68)^62^ and the ABF function within the R package “coloc” (version 5.1.0)^63^. For eQTL analyses, these were performed in all specifically enriched tissues (via LDSC-SEG)^64^, as well as the cross-tissue meta-analysed GTEx eQTL data v.7^65^ and data from the eQTLGen^66^ and Brain-eMeta^67^ studies. Lastly, genes residing within each locus underwent prioritisation using the gene-level polygenic priority score (PoPS) method^68^. Causal candidate genes were then prioritised by overlaying all of the above information and scoring the strength of evidence observed. For further details about the specific application of this method, see Kentistou *et al*.^19^.

This was conducted on each of the timepoint-specific MoBa and gSEM GWAS. To prioritise likely causal genes at each GWAS signal, we prioritised the highest-scoring gene each locus, if supported by at least 2 concordant predictors from the above.

### Identification of signals and genes across GWAS

After identifying conditionally independent GWAS signals separately for each of the 11 child BMI timepoints in the MoBa cohort (as described above), we combined these to derive a single list of independent signals across childhood. To do this, we first took all the overlapping timepoint-specific signals and calculated an LD *R*^2^ matrix. We then retained all LD-independent signals (*R*^2^<0.05), while prioritising the later MoBa timepoints (i.e. 8 years, over 7 years, over 5 years, etc.). If a signal at 1 timepoint was correlated (*R*^2^>0.5) with a signal at a different timepoint, the SNP and timepoint with the smaller *P*-value was retained.

We further combined signals from the above across-timepoint list with independent signals from the gSEM GWAS. This was done by retaining the MoBa signals and adding any uncorrelated (*R*^2^<0.05) gSEM signals (excluding heterogenous gSEM signals with a Q-index, *P*_Q,_<5×10^-8^). Gene prioritisation for each signal was derived from the G2G data in the most significant timepoint for MoBa BMI signals, and in the gSEM G2G analysis for gSEM signals.

### Clustering of variants into trajectories

Hierarchical clustering was performed to identify trajectories of SNPs with similar effects throughout childhood. We considered the effect of the variants across the MoBa time points (including birthweight, even though this was not used for discovery), as well as SAC10 from UK Biobank and the gSEM. To assess the effect sizes’ trajectories rather than the absolute difference in effect sizes, correlation was used as dissimilarity metric. This dissimilarity metric was calculated by *(1-p)/2*, where *p* is the correlation between 2 SNPs’ effect estimates in the twelve GWAS. Complete linkage was used to determine the distance between 2 clusters. Prior work^16^, using supervised clustering, had suggested 4 trajectories and using a cut-point resulting in 4 trajectories recapitulated the patterns seen there. The homogeneity of trajectories was confirmed using a heatmap where the consistency of effect estimates across trajectories was compared.

### Mendelian Randomisation

We performed inverse weighted 2-sample Mendelian randomisation (MR)^69^ for Type 2 diabetes and several anthropometric phenotypes of relevance to metabolic disease risk. We used published data for Type 2 Diabetes^70^ and GWASs performed on UK Biobank data for body fat percentage, fat mass, lean mass, and waist-to-hip ratio (WHR). We performed MR analyses separately for the SNPs in each early BMI trajectory. To optimise exposure estimates, for each early BMI trajectory we weighted each SNP by its effect estimate on BMI at a representative MoBa timepoint: 6 months for the transient SNPs, 1 year for the early rise SNPs, 8 years for the late rise SNPs. In the case of the birth-related SNP, as this has a “U”-shaped distribution, we performed the analysis weighting by both the effects for BMI at birth and at 8 years. To investigate if the trajectories conferred any risk beyond that of increased adiposity in adulthood, we additionally performed a multivariate MR accounting for the SNP effects on adult BMI.

### Analysis of Parent-of-Origin effect and Imprinting

We identified and extracted transmitted and non-transmitted alleles for each SNP within parent-offspring trios, classifying them into maternally transmitted (mt), paternally transmitted (pt), maternally non-transmitted (mnt), and paternally non-transmitted (pnt) alleles. We employed linear regression models to evaluate the association between transmitted and non-transmitted alleles and child BMI at the most significant timepoint in the MoBa cohort, adjusting for covariates such as sex, birth cohort, and the top 10 PCs to control for population structure. We excluded pnt alleles from the model due to both the smaller sample size and the reduced biological relevance of these alleles. We estimated the beta model parameters for mt and pt alleles, as well as their differences (β_mt_ − β_pt_) to quantify PoE. We assessed their statistical significance using a general linear hypothesis test for the null hypothesis (β_mt_ − β_pt_ = 0) using a t-distribution. We investigated SNPs located near known imprinted genes (within 100kb)^71^ to determine if these regions exhibited PoE. Replication was conducted in up to 123,716 unrelated individuals from the UK Biobank for SAC10 and adult BMI traits using the PoE inference method described in Hofmeister *et al.*^72^.

### Biological Pathway Enrichment Analysis

We performed pathway enrichment analysis using g:Profiler (via the R client “gprofiler2”, version 0.2.242). We used pathways from the GO, KEGG and REACTOME databases (accessed on 07/05/2024) and restricted the analysis to pathways with 1000 genes or fewer, reasoning that these are more biologically specific. Pathway enrichment analyses were performed using the combined set of variants from either MoBa or the gSEM. Pathways with a Bonferroni corrected *P*<0.05 were considered to be enriched.

As many reference pathways are based on overlapping data, we aggregated the childhood adiposity-associated pathways based on any shared intersect genes. We used a “complete” clustering algorithm, and a custom distance calculated as [1 minus the proportion of the overlap between any 2 pathways relative to the pathway with the smaller size]. Thus, between 2 given pathways, a value of 0 indicates that all intersecting genes in the pathway with fewer genes are also present in the other pathway. To define clusters, we chose an arbitrary cut-off value of 0.5, which indicates that pairs of pathways share 50% or more genes.

### Child-adult effect comparisons

We used GWAS data on adult BMI from the GIANT consortium^20^ to explore which of our identified GWAS signals exert larger effects on adiposity in childhood compared to adulthood. We categorised child adiposity signals which had no evidence of association with adult BMI (*P*>0.05) or a nominal association with adult BMI (*P*<0.05) but with the opposite directions of effect on childhood and adulthood BMI. We also rescaled the gSEM common factor effect estimates to a zBMI scale, similar to the cross-sectional MoBa GWAS and adult BMI GWAS. To do this, we regressed the effect estimates of the 152 SNPs discovered in the MoBa-GWASs against their effect estimates on the gSEM common factor and used the slope of this association (0.5365501) as a scaling factor (i.e. gSEM common factor effect estimate / 0.5365501). We then performed a look-up of all independent child adiposity GWAS signals in the adult BMI GWAS, using the best available proxy (with *R*^2^>=0.4) for any child signal unavailable in the adult BMI GWAS. We aligned all SNPs by the child adiposity-increasing allele, and calculated the difference in effect estimates between childhood and adulthood (i.e. β_child_-β_adult_). We considered the top 5% of signals in the distribution of these effect differences to have disproportionately larger effects on child BMI than adult BMI.

### Integration of human hypothalamic cell data

We used the Human HYPOMAP data from Tadross *et al.*^32^ to identify hypothalamic cell populations which are specifically enriched for genes harbouring either childhood or adult adiposity associated variants. For adult adiposity, we used GWAS data from the GIANT BMI meta-analysis^20^ (Nmax=806,834) and for child adiposity we used the filtered gSEM summary statistics, as described above. As these were genome-wide data, alongside filtering for signals exhibiting significant heterogeneity (pQ<5×10^-8^), we also removed all other variants in even weak LD with these Q-SNPs (R^2^>0.05). We then used cell type specificity matrices, generated viaCELLEX (https://github.com/perslab/CELLEX) as described in Tadross *et al.*^32^, in CELL-type Expression-specific integration for Complex Traits (CELLECT) with Multi-marker Analysis of GenoMic Annotation (MAGMA). For each trait, CELLECT-MAGMA (v1.3.0) was run with default parameters across the 452 hypothalamic cell populations with a multiple-test corrected significance threshold of *P*<0.05/452.

To identify childhood adiposity genes that contributed strongly to cell-type classification (‘informative marker genes’), we defined genes that were both i) a high-confidence childhood adiposity genes identified through G2G, and ii) in the top 100 genes (equivalent to the ∼99.5^th^ percentile) for cell-type specificity within each identified cell-cluster. This approach was used to highlight childhood adiposity genes that are specifically expressed in the child-adiposity enriched ARC or MAM cell populations (**Supplementary Data 21**).

### Gene burden testing of rare variants in whole-genome sequence data

Whole-genome sequencing (WGS) data in up to 490,640 participants from UK Biobank was accessed and processed as described previously^73^. Briefly, 1,037,556,156 SNPs and 101,188,713 indels were called using Graphtyper^74^. Multi-allelic sites were split and normalized in Hail v0.2^75^ and variants with on low QC metrics or high missingness were filtered out. Variants were initially annotated using the Ensembl Variant Effect Predictor (VEP^76^; v108.2) to identify missense and protein-truncating variants (PTVs; defined as any variant with stop-gained, splice acceptor, and splice donor consequence), with the LOFTEE^77^ plugin for the identification of high-(HC) and low-confidence (LC) PTVs. We also used REVEL^34^ (rare exome variant ensemble learner) scores to annotate missense variants. We tested associations with 2 variant collapsing masks. One mask comprised of rare (MAF<0.1%) HC PTVs within each gene. The other mask comprised of rare (MAF<0.1%) predicted deleterious missense variants (defined by REVEL score ≥0.7).

We tested 2 phenotypic outcomes; size as child, age 10 (SAC10), as described above, and adult BMI (kg/m^2^) from field 23104 and BOLT-LMM (v2.4.1)^51^. Gene burden tests were performed by generating a dummy genotype file in which each gene-mask combination was coded as a single variant. We then coded individuals with 1+ qualifying variants for each mask within a gene as heterozygous. When testing for association against the 2 outcomes in BOLT-LMM, we used the ‘lmmInfOnly’ flag and the following covariates; age, age2, sex, age*sex, the first 20 principal components, and the WGS-release batch. We included all samples without restricting their ancestries to maximise the sample size and only looked at genes with at least 100 carriers of qualifying variants. Only samples who withdrew consent or had missing phenotypes and covariates were excluded and this filtering resulted in 481,137 and 479,615 samples remaining for BMI and SAC10, respectively.

### Related trait WGS associations

For all identified gene burden associations with SAC10, we performed similar gene burden association testing for predefined relevant obesity-related phenotypes. We used genetic data from the previous release of whole-exome sequencing data in UK Biobank, which was conducted in up to 450,000 participants.

We selected 8 phenotypes, inclusive of SAC10 and adult BMI, defined as follows. Body-fat-percentage was extracted using data from UK Biobank field 23099. For all phenotypes, data from subsequent visits were used, if missing for a given instance. Comparative height at age 10 (HAC10) was recoded to have individuals who in field 1697 reported being ‘Shorter’ as 0, ‘About average’ as 1, and ‘Taller’ as 2. Individuals who reported ‘Do not know’ or ‘Prefer not to answer’ were set to ‘NA’. Following variable recoding, this phenotype was run as a continuous trait. WHR adjusted for BMI: WHR was calculated using data from fields 48 and 49. Individuals who were who were 4 or more standard deviations from the mean were excluded after the initial calculation and prior to adjusting with BMI (field 21001). Paired BMI and WHR data from subsequent visits were used, if missing for a given instance. T2D was derived as described in^56^ and using data from fields 4041, 10844, 2443, 6177, 6153, 20002, 20003, 41202, 41204, 40001 and 40002. A lean mass index was calculated similarly to BMI; but using whole body fat-free mass, derived from the bioelectrical impedance analysis field 23101 as the numerator, and height squared in centimetres from field 50 as the denominator. Again, data from subsequent visits were used, if missing for a given instance. Average hand grip strength was derived from fields 46 and 47. Data from subsequent visits were used, if missing for a given instance. The multiple testing threshold was set at *P*<0.05/(3×6×8) ∼ 3.5×10^-4^, after correcting for the above 8 queried phenotypes, 6 identified genes, and 3 variant collapsing masks.

We conducted gene burden tests against these 8 outcomes, by combining the effects of all rare (MAF<0.1%) variants with predicted deleterious functional consequences as described in Gardner *et al*.^78^, using data in up to 419,692 individuals of European ancestry from UK Biobank^79^. We collapsed rare variants into 3 masks: HC PTVs, missense variants with REVEL score ≥0.7, and missense variants with CADD score ≥25. While this dataset was very similar to the WGS dataset used in the discovery analysis, there were some differences stemming from the difference in cohort size, sequencing technologies, variant annotation and prioritisation strategies as outlined above.

## Acknowledgements

This work was supported by grants (to S.J) from Helse Vest’s Open Research Grant (#912250 and F-12144), the Novo Nordisk Foundation (NNF20OC0063872) and the Research Council of Norway (#315599). J.S. and S.J. was supported by the Meltzer Research Fund. K.A.K., L.R.K. Y.Z., F.R.D., J.R.B.P., and K.K.O. were supported by the UK Medical Research Council (MC_UU_00006/2) and the NIHR Cambridge Biomedical Research Centre. M.V. was supported by the Research Council of Norway (#301178), the European Research Council (#101171420), and the University of Bergen. J.A.T., B.Y.H.L. and G.S.H.Y. are supported by BBSRC Project Grant (BB/S017593/1) and the MRC Metabolic Diseases Unit (MC_UU_00039/1). G.K.C.D. is supported by a BBSRC iCASE studentship co-financed by Novo Nordisk. LS and JCB are supported by Novo Nordisk A/S, the German National Diabetes Centre (DZD) and European Research Council (ERC) grant (SYNEME: 742106). S.O is supported by the MRC Metabolic Diseases Unit (MC_UU_00039/1) and the NIHR Cambridge Biomedical Research Centre (NIHR203312). S.L. is supported by a Wellcome Trust Clinical PhD Fellowship (225479/Z/22)

MoBa: We thank the Norwegian Institute of Public Health (NIPH) for generating high-quality genomic data. This research is part of the HARVEST collaboration, supported by the Research Council of Norway (#229624). We also thank the NORMENT Centre for providing genotype data, funded by the Research Council of Norway (#223273), South East Norway Health Authorities and Stiftelsen Kristian Gerhard Jebsen, and in collaboration with deCODE Genetics. We further thank the Center for Diabetes Research, the University of Bergen for providing genotype data and performing quality control and imputation of the data funded by the ERC AdG project SELECTionPREDISPOSED, Stiftelsen Kristian Gerhard Jebsen, Trond Mohn Foundation, the Research Council of Norway, the Novo Nordisk Foundation, the University of Bergen, and the Western Norway Health Authorities. We thank the MoBaPsychGen team, led by Elizabeth Corfield, for providing quality controlled genotype data, supported by funding from the South-Eastern Norway Regional Health Authority (#2021045; #2020022; #2022083; 2018058).

We are grateful to all the families in Norway who are taking part in the ongoing MoBa cohort study. All analyses in the MoBa cohort were performed using digital labs in HUNT Cloud at the Norwegian University of Science and Technology, Trondheim, Norway. We are grateful for outstanding support from the HUNT Cloud community.

ALSPAC: We are extremely grateful to all the families who took part in the ALSPAC cohort study, the midwives for their help in recruiting them, and the whole ALSPAC team, which includes interviewers, computer and laboratory technicians, clerical workers, research scientists, volunteers, managers, receptionists and nurses. The UK Medical Research Council and Wellcome (217065/Z/19/Z) and the University of Bristol provide core support for ALSPAC. This publication is the work of the authors and S.J. will serve as guarantor for the contents of this paper. A comprehensive list of funding is available on the ALSPAC website (http://www.bristol.ac.uk/alspac/external/documents/grant-acknowledgements.pdf); this research was specifically funded by Wellcome Trust and MRC (core) (076467/Z/05/Z). ALSPAC GWAS data was generated by Sample Logistics and Genotyping Facilities at Wellcome Sanger Institute and LabCorp (Laboratory Corporation of America) using support from 23andMe.

## Ethics declarations

MoBa: Informed consent was obtained from all study participants. The administrative board of the Norwegian Mother, Father and Child Cohort Study led by the Norwegian Institute of Public Health approved the study protocol. The establishment of MoBa and initial data collection was based on a license from the Norwegian Data Protection Agency and approval from The Regional Committee for Medical Research Ethics. The MoBa cohort is currently regulated by the Norwegian Health Registry Act. The study was approved by The Regional Committee for Medical Research Ethics (#2012/67).

UK Biobank: Informed consent was obtained from all study participants. The study has approval from the North West Multicentre Research Ethics Committee as a Research Tissue Bank.

ALSPAC: Ethical approval for the study was obtained from the ALSPAC Ethics and Law Committee and the Local Research Ethics Committees. Informed consent for the use of data collected via questionnaires and clinics was obtained from participants following the recommendations of the ALSPAC Ethics and Law Committee at the time. Consent for biological samples has been collected in accordance with the Human Tissue Act (2004).

## Author information

### Author contribution

**K.A.K., J.S., F.R.D., M.V., J.R.B.P., K.K.O., and S.J.** wrote the manuscript. **K.A.K., J.S., R.K., L.R.K., R.J.H., A.E.L., N.F.B., F.R.D., and M.V.** performed data analyses and interpretation. **J.A.T., L.S., G.K.C.D., J.C.B., B.Y.H.L., and G.S.H.Y.** contributed with data, analysis and interpretation of Hypomap. **Y.Z., J.L., A.C., Y.L., and J.D.** contributed with data, analysis and interpretation of WGS. **S.L., L.C., A.H., O.A.A., E.B., S.O.R., P.R.N and Z.K.** contributed with additional data analyses and interpretation. **F.R.D., M.V., J.R.B.P., K.K.O., and S.J.** supervised the work. All authors critically reviewed and approved the final version of the manuscript.

## Data availability

Data from the Norwegian Mother, Father and Child Cohort Study used in this study are managed by the Norwegian Institute of public health and can be made available to researchers, provided approval from the Regional Committees for Medical and Health Research Ethics (REC), compliance with the EU General Data Protection Regulation (GDPR) and approval from the data owners. The consent given by the participants does not open for storage of data on an individual level in repositories or journals. Researchers who want access to data sets for replication should apply through helsedata.no. Access to data sets requires approval from The Regional Committee for Medical and Health Research Ethics in Norway and an agreement with MoBa.

The UK Biobank phenotype and genetic data described here are publicly available to registered researchers through the UKB data access protocol. Information about registration for access to the data is available at: www.ukbiobank.ac.uk/enable-your-research/apply-for-access. This work was conducted using UK Biobank data, application number 9905.

All bona fide researchers can apply to use ALSPAC data for health-related research that is in the public interest. Information regarding the ALSPAC cohort and data access is available at www.bristol.ac.uk/alspac/researchers/our-data.

GWAS summary statistics can be obtained from the authors and will be made publicly available after peer review.

