## Supplementary Information for "The role of common and rare genetic variation on adiposity across childhood"

**Page 2. Supplementary Figure 1** | Performance of time-stratified genomic PRSs (PRS_GW) based on MoBa discovery to predict childhood BMI in ALSPAC.

**Page 3. Supplementary Figure 2** | Scatterplots comparing common factor and individual trait associations (Z-scores) for the genomic SEM GWAS signals.

**Page 4. Supplementary Figure 3** | Performance of the childhood adiposity common factor PGS (PGS_CommonFactor) to predict a) childhood BMI and b) childhood fat percentage in ALSPAC.

**Page 5. Supplementary Figure 4** | Performance of trajectory-specific PGS to predict childhood BMI in ALSPAC

**Page 6. Supplementary Figure 5** | Enriched biological pathways.

**Page 7. Supplementary Figure 6** | Neuronal cell populations are enriched for genes linked to childhood adiposity variation in the general population.

**Page 8. Supplementary Figure 7** | Sensitivity analysis for WGS identified genes for childhood adiposity in UK Biobank WES data.

**Page 9. Generation of genomic SEM common factor models**

### Supplementary Figures

**Supplementary Figure 1 | Performance of time-stratified genomic PRSs (PRS_GW) based on MoBa discovery to predict childhood BMI in ALSPAC**. For comparison, performance of a PGS based on adult BMI GWAS data (Yengo 2018) if also shown. Further data can be found in Supplementary Table 6.

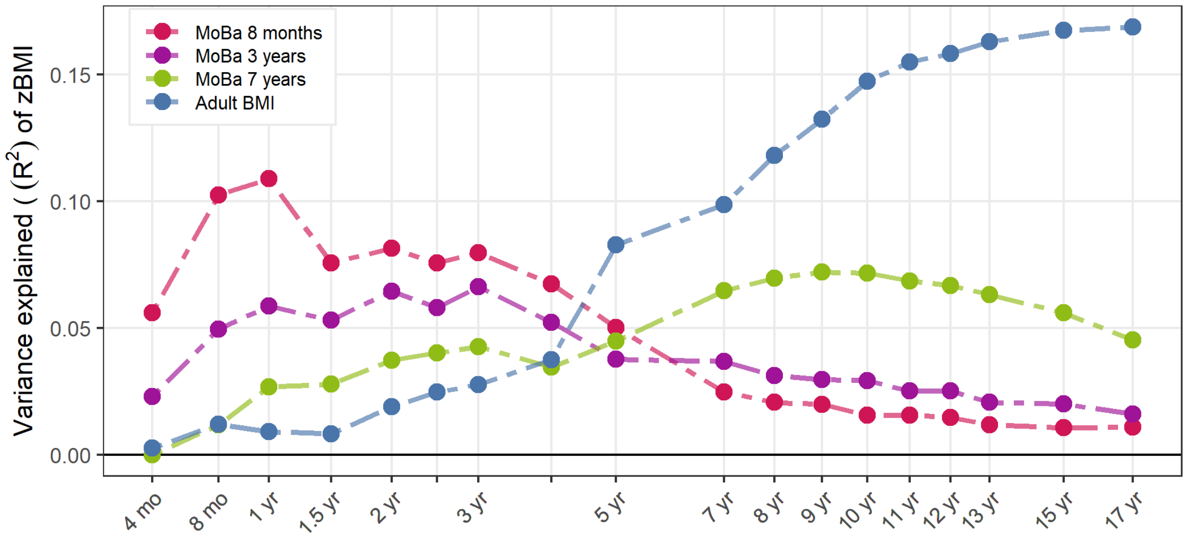

**Supplementary Figure 2 | Scatterplots comparing common factor and individual trait associations (Z-scores) for the genomic SEM GWAS signals.** Heterogeneous signals ($P_Q<5\times10^{-8}$) are highlighted in orange; all of these except for rs663129 (which maps to *MC4R*) were excluded in subsequently genetic analyses. AAM=age at menarche, MOBA8=childhood BMI age 8, SAC10=comparative body size age 10. Further data can be found in **Supplementary Table 8**.

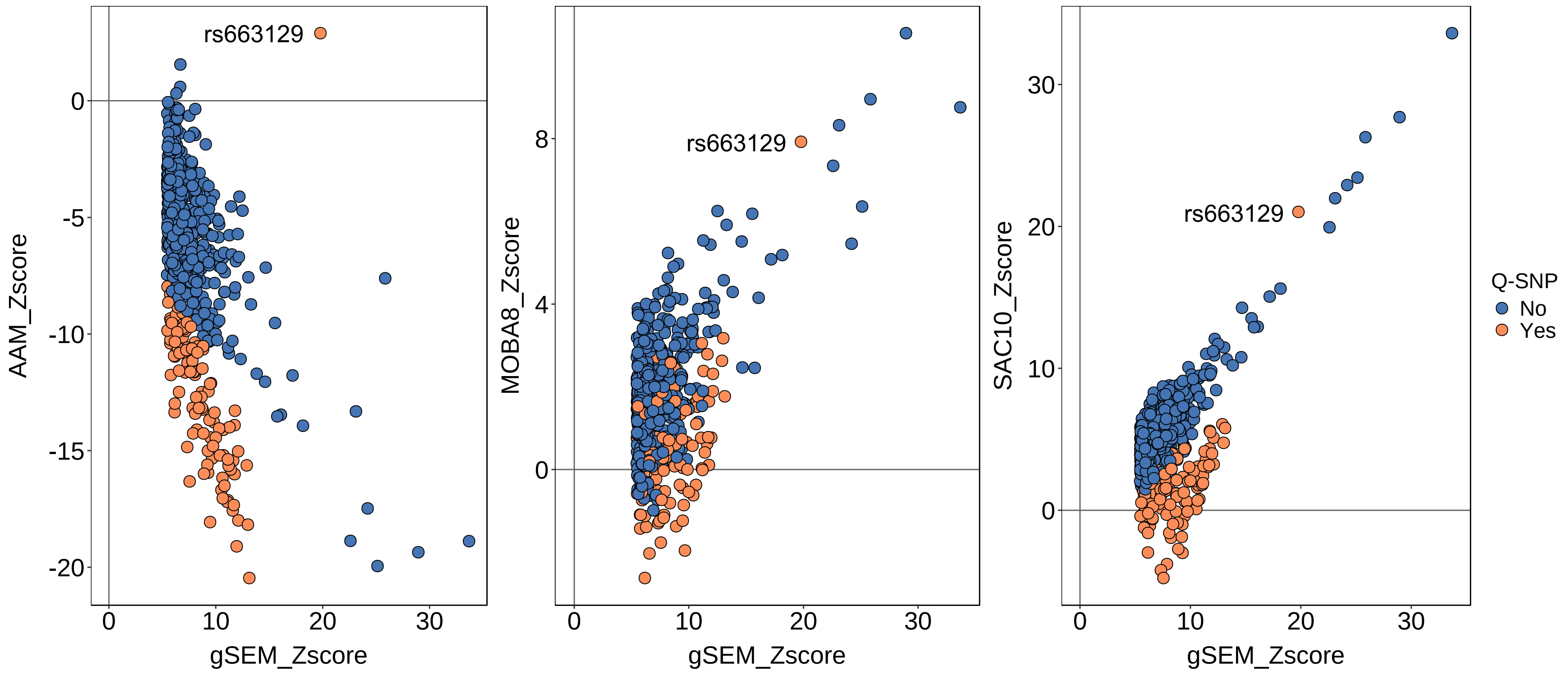

**Supplementary Figure 3 | Performance of the childhood adiposity common factor PGS (PGS_CommonFactor) to predict a) childhood BMI and b) childhood fat percentage in ALSPAC**. For comparison, performance of a PGS based on recalled size at age 10 (SAC10), and a PGS based on adult BMI GWAS data (Yengo 2018) are also shown. Further data can be found in Supplementary Table 7.

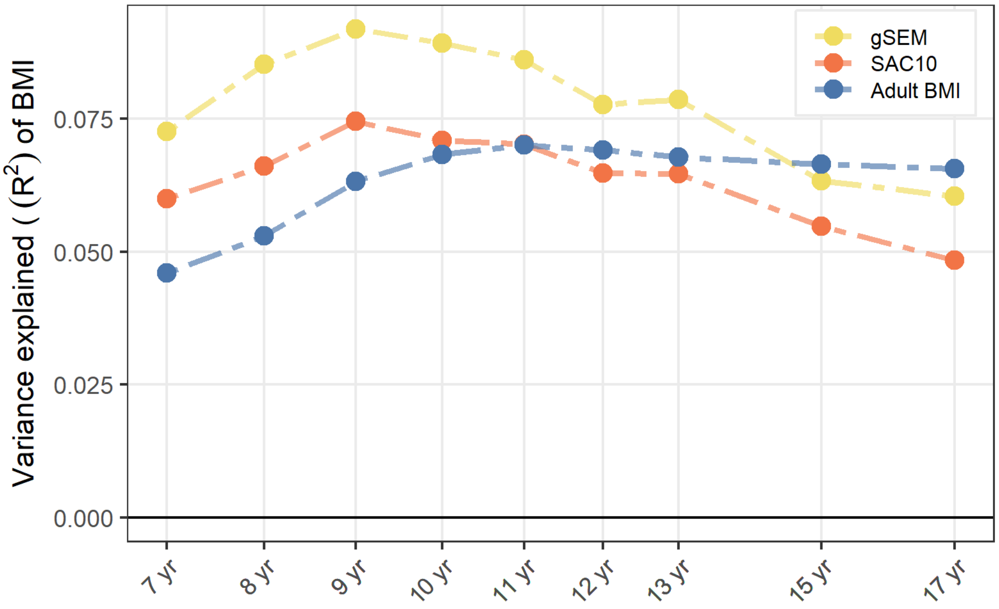


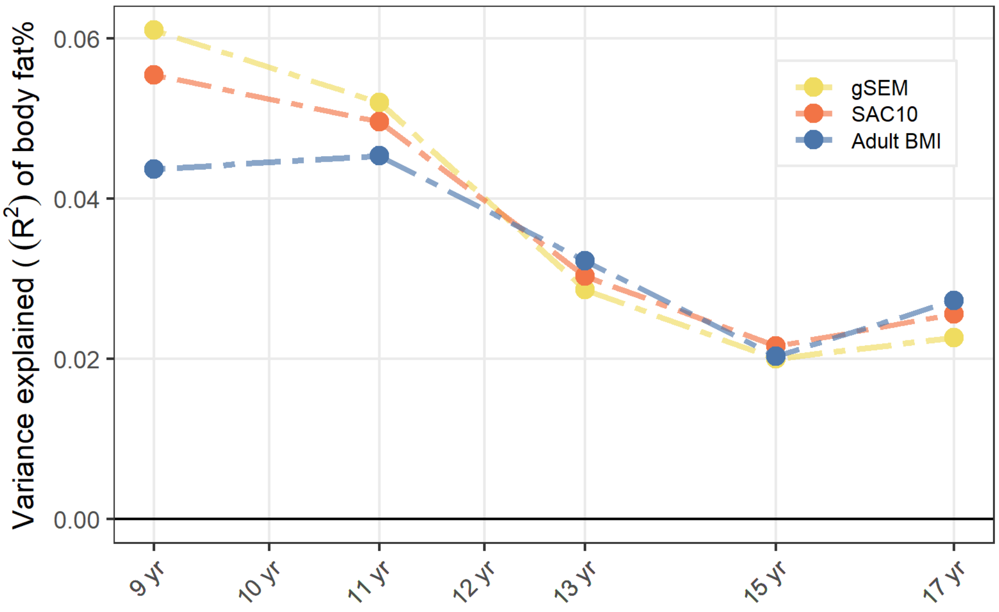

**Supplementary Figure 4 | Performance of trajectory-specific PGS to predict childhood BMI in ALSPAC.** Underlying data can be found in Supplementary Data 11.

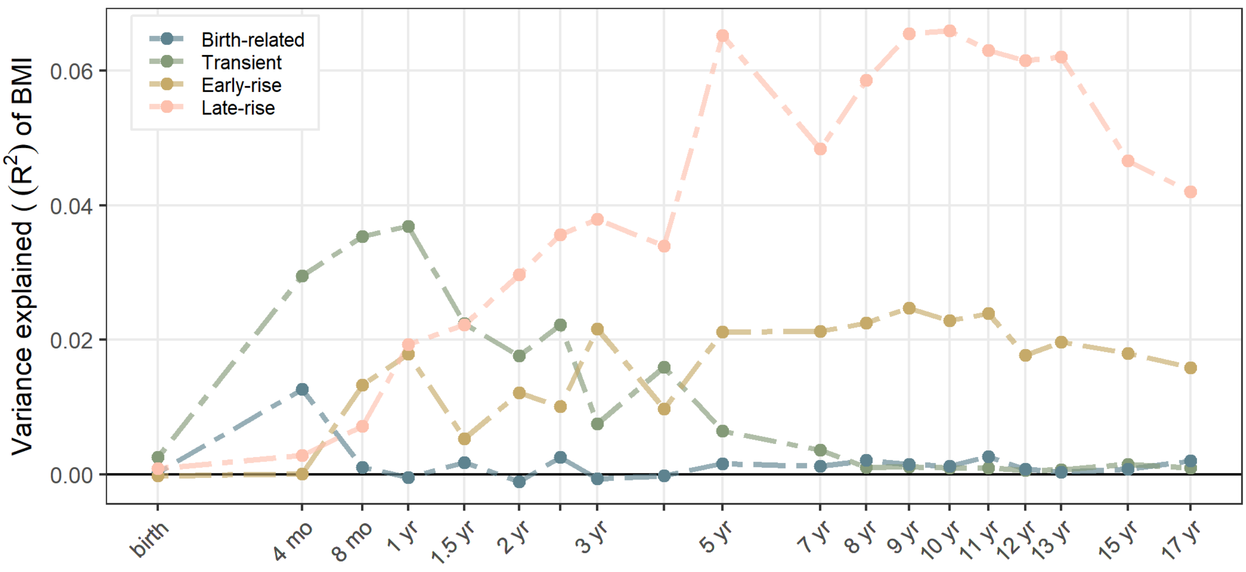

**Supplementary Figure 5 | Enriched biological pathways.** a) Dendrogram of pathways enriched for childhood adiposity association signals, analysed in gProfiler. Clustering is scaled by our custom distance metric (see **Methods**). Pathway names are coloured if also enriched for association signals in each of the four trajectories. b) Numbers of overlapping pathway between trajectories. c) Trajectory-specific pathway groups and their individual pathways, along with the genes driving pathway enrichment. More details can be found in **Supplementary tables 17** and **18**.

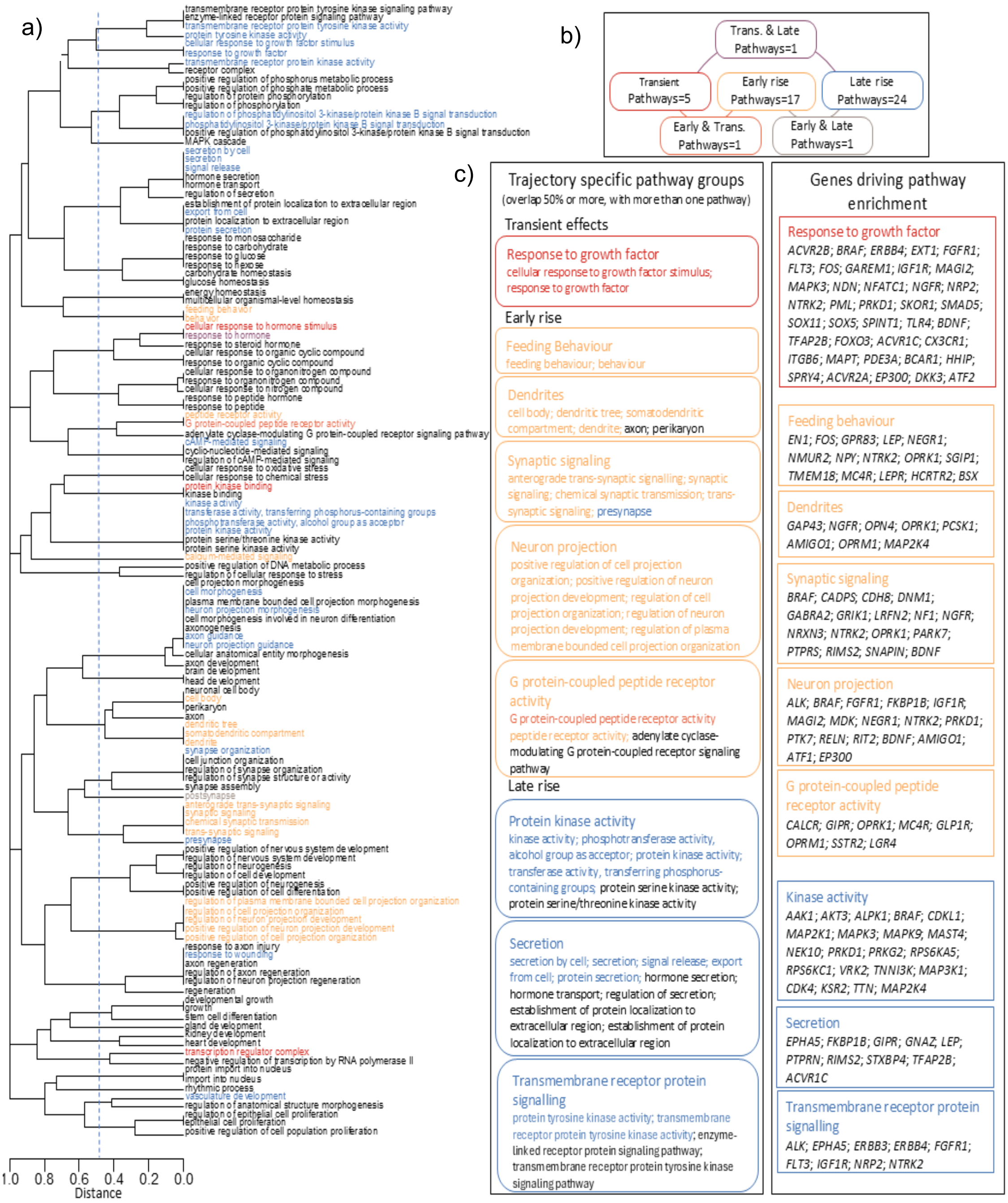

**Supplementary Figure 6 | Neuronal cell populations are enriched for genes linked to childhood adiposity variation in the general population.** Prioritisation of 452 human hypothalamic cell types identified 251 cell types as significantly enriched for associations in the filtered gSEM GWAS. Each point represents a cell type and cell types were grouped (y-axis) by neuronal or non-neuronal cell types and coloured based on specific regions assigned via marker-gene expression. The dashed line indicates a Bonferroni-corrected significance threshold, $P<0.05/452$. Further data can be found in **Supplementary Table 20**.

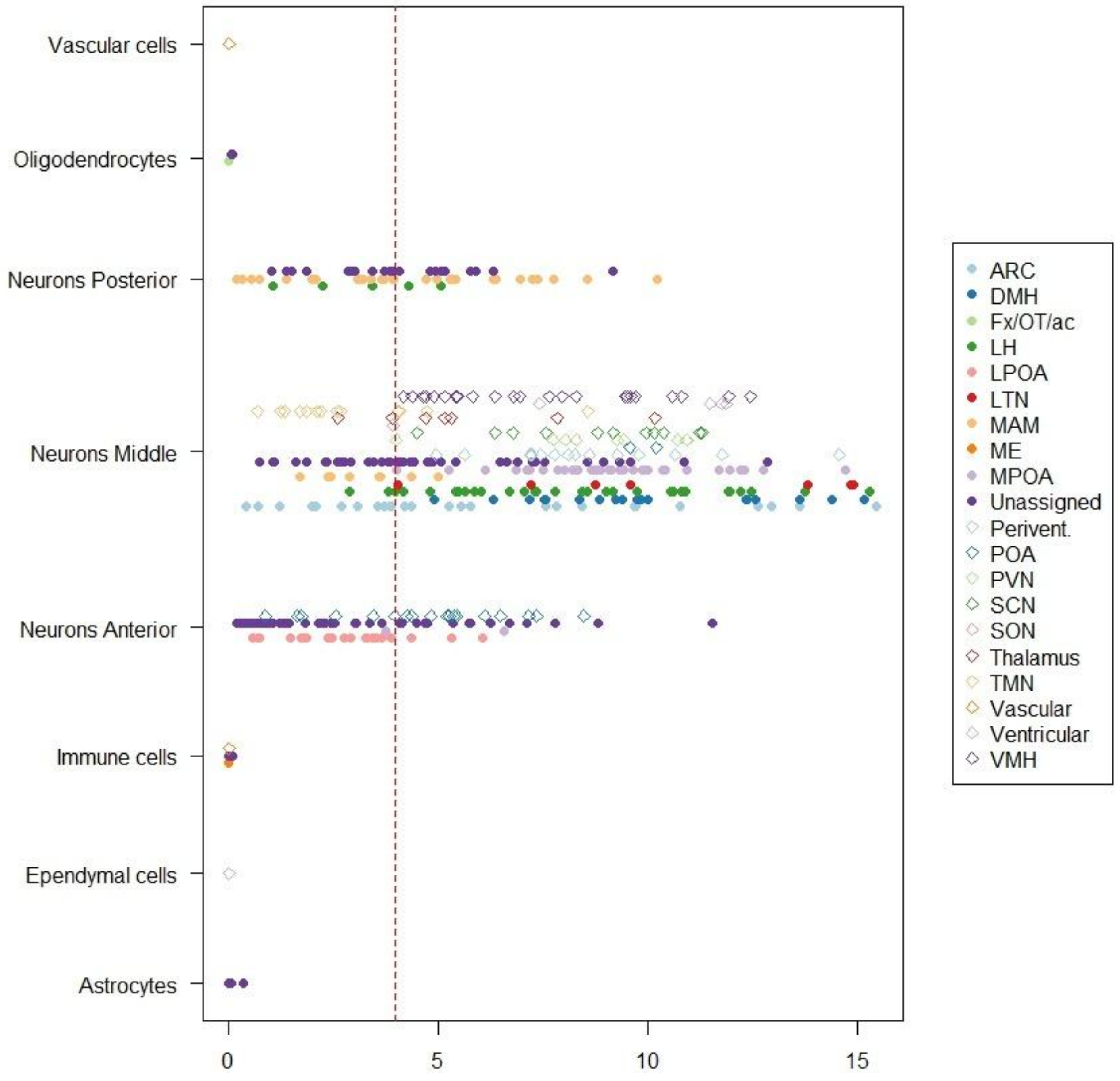

**Supplementary Figure 7 | Sensitivity analysis for WGS identified genes for childhood adiposity in UK Biobank WES data.** Gene burden test effect estimates from BOLT LMM on SAC10 and adult BMI for the six genes identified in the UK Biobank WGS analysis of rare deleterious variants. For comparison, similar effect estimates are shown in the smaller UK Biobank WES dataset. Point opacity indicates at least nominal association (*P*<0.05, two-sided). Point shapes indicate the variant collapsing mask and colours indicate the source dataset. Error bars indicate 95% confidence intervals. Further data can be found in **Supplementary Tables 22 and 23**.

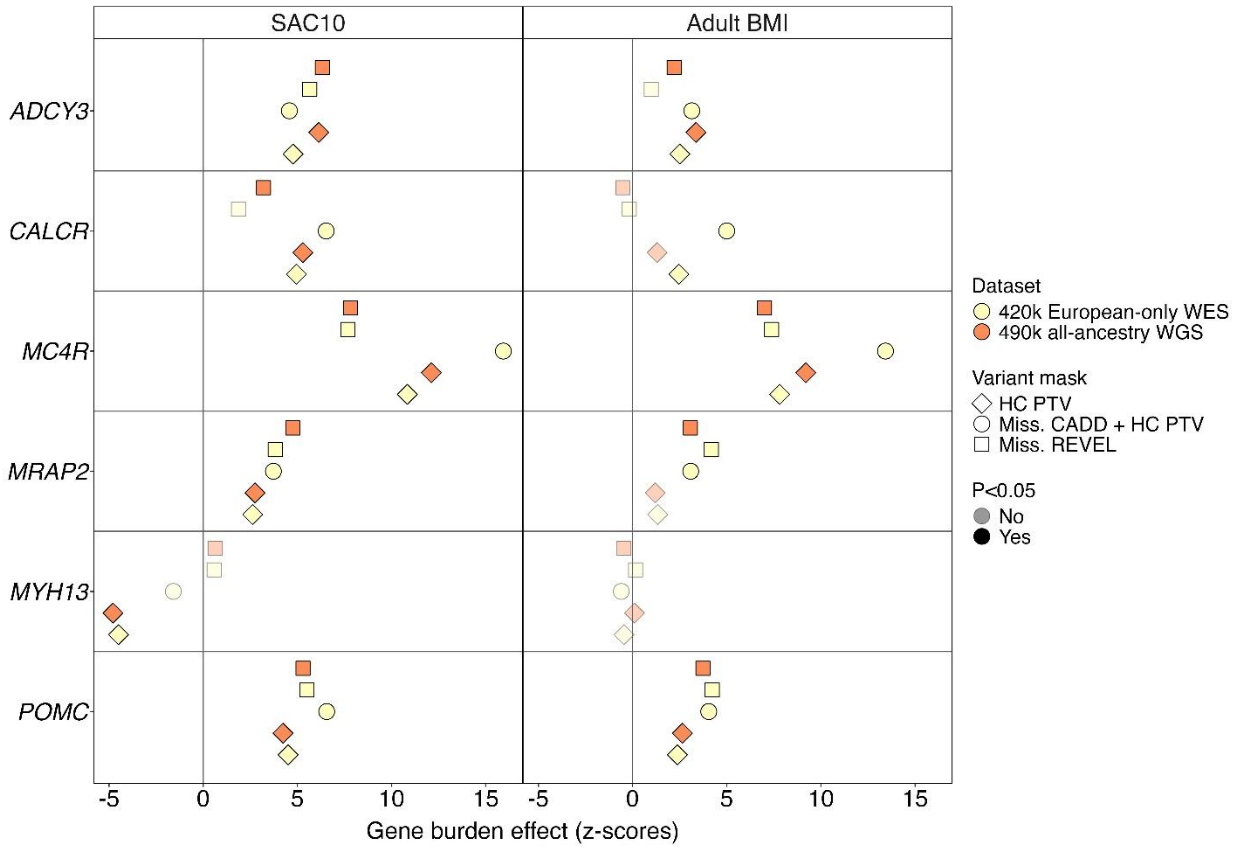

### Generation of genomic SEM common factor models

Correlations between individual traits: We observed a strong positive genetic correlation between SAC10 and childhood BMI (MOBA7: $r_g$=0.88; MOBA8: $r_g$=0.85) consistent with previous reports. We also observed a moderately strong inverse genetic correlation between each childhood adiposity trait and AAM ($r_g$: -0.36 to -0.39), similar to the reported genetic correlation between AAM and adult BMI (**Figure 1**).

***Figure 1*** **|** *Heatmap of genetic correlations among AAM (age at menarche), SAC10 (comparative body size age 10), MOBA7 (childhood BMI age 7) and MOBA8 (childhood BMI age 8). Correlations were estimated using multivariate LDSC.*

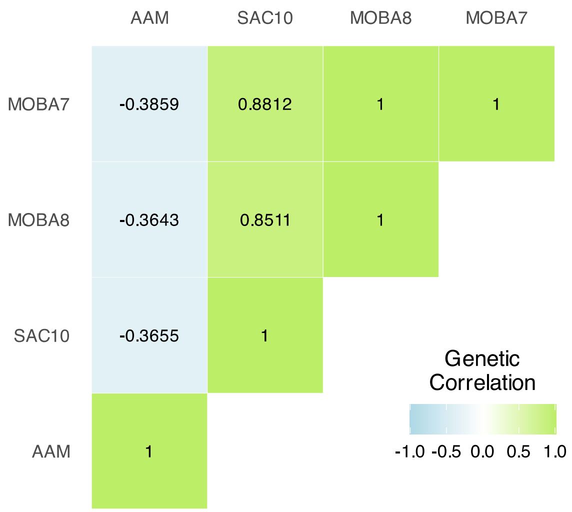


These genetic correlations supported the rationale to run our pre-specified genomic SEM models assuming the presence of a single "childhood adiposity" common factor:

1. AAM, SAC10, MOBA7
2. AAM, SAC10, MOBA8
3. AAM, SAC10, MOBA7, MOBA8.
4. SAC10, MOBA7, MOBA8.

Genomic SEM model fit: The model fit for the first three models was good**.** Model 4 generated a 'Heywood case' where the standardised factor loading from MOBA7 exceeded 1 and thus also had a negative residual variance. If we imposed additional model constraints to keep the residual variance >0, the degrees of freedom for Model 4 dropped to 0, and no model fit indices could be calculated.

Genomic SEM factor loadings: Model results showed strong positive loadings for all adiposity traits across all four models (0.87-1.00) (**Figure 2**). In Models 1, 3, and 4, MOBA7 showed the strongest factor loading (0.96 to 0.99). In Model 2, SAC10 and MOBA8 showed similarly strong factor loading (both 0.92). AAM showed moderate negative loadings (-0.40) on Models 1,2 and 3, consistent with the genetic correlations between individual traits (**Figure 1**).

***Figure 2 |*** *Common factor analysis of the genetic "childhood adiposity" common factor (F1) for a) Model 1, b) Model 2, c) Model 3 and d) Model 4. Standardised indicator loading estimates of each individual trait on the common factor are displayed (AAM=age at menarche, SAC10=comparative body size age 10, MOBA7=childhood BMI age 7, MOBA8=childhood BMI age 8). The u variables indicate residual variances in each trait.*

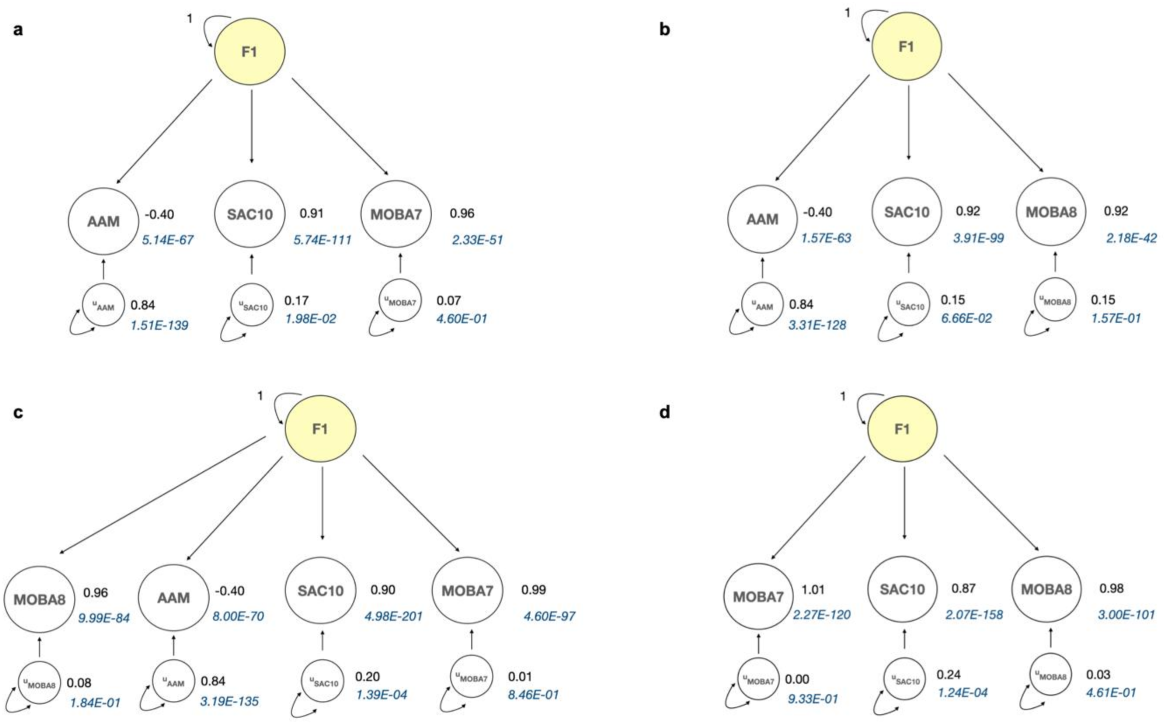

GWAS signals for each genomic SEM common factor: We incorporated genome-wide SNP effects into the genetic covariance matrix and the sampling covariance matrix to estimate the effect of each SNP on the "childhood adiposity" common factor in each model.

- Model 1 identified 510 independent GWAS signals ($p<5x10^{-8}$) for its "childhood adiposity" common factor. Of the 510 loci, 66 (13%) did not reach genome-wide significance for association with any contributing individual trait (**Figure 3a**).
- Model 2 identified 643 independent GWAS signals. Of these, 135 (21%) did not reach genome-wide significance for association with any contributing individual trait (**Figure 3b**).
- Model 3 identified 539 independent GWAS signals. Of these 106 (20%) did not reach genome-wide significance for association with any contributing individual trait (**Figure 3c**).
- Model 4 identified 258 independent GWAS signals. Of these 35 (14%) did not reach genome-wide significance for association with any contributing individual trait (**Figure 3d**).

Model 2 produced the most GWAS signals and the largest effective sample size. This is because the trait with the highest factor loading in this model (SAC10, slightly larger than MOBA8) had a much larger sample size and thus better-powered effect estimates than MOBA7, which had the highest factor loading in Models 1, 3 and 4.

***Figure 3*** **|** *Manhattan plots showing GWAS results for the "childhood adiposity" common factors generated by Models 1-4 (a-d). Red horizontal lines indicate genome-wide significance ($p<5\times10^{-8}$). Independent genome-wide significant loci are highlighted in red.*

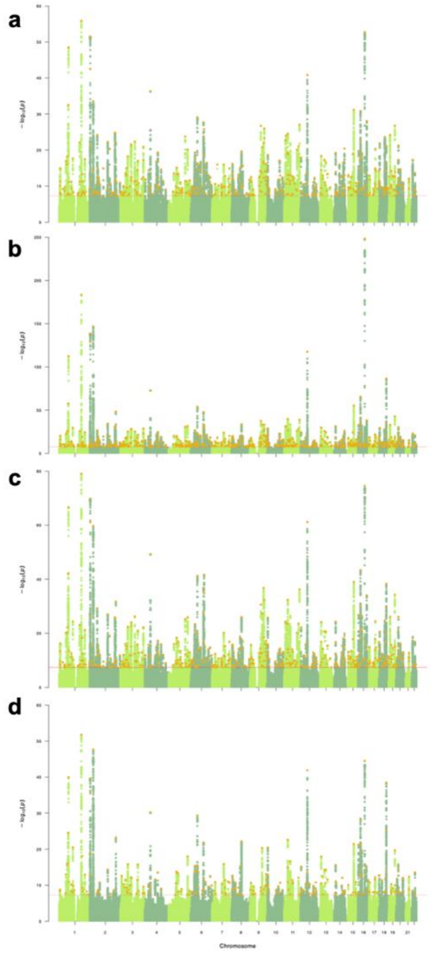

QQ plots of GWAS results for the common factors from all models showed slightly higher genomic inflation factors ($\lambda_{GC}$ =1.69-2.04) compared to those for each individual trait (AAM $\lambda_{GC}$ =1.72, SAC10 $\lambda_{GC}$ =1.43, MOBA7 $\lambda_{GC}$ =1.14, MOBA8 $\lambda_{GC}$ =1.12) (**Figure 4**). Reassuringly, all LDSC intercepts were close to one, indicating that $\lambda_{GC}$ values reflect polygenicity rather than uncontrolled inflation.

***Figure 4* |** *Quantile-quantile (QQ) plots of p-values from GWAS for each "childhood adiposity" common factor generated by Models 1-4 (a-d). Expected -log$_{10}$ p-values are those expected under the null hypothesis.*

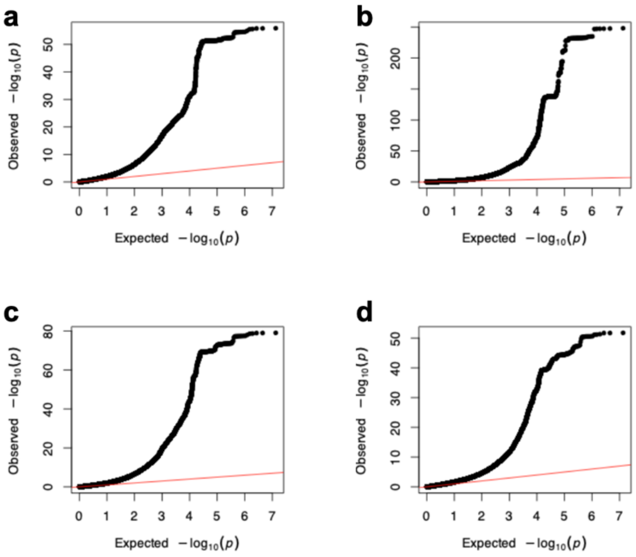

Furthermore, lead SNPs for each “childhood adiposity” common factor in Models 1-4, but which did not show genome-wide significant association ($P>5x10^{-8}$) with any individual contributing trait, showed near but sub-threshold genome-wide significant association with individual traits (**Figure 5**). This confirms that genomic SEM was unlikely to generate artefactual SNP associations, e.g. due to collider effects.

***Figure 5*** **|** *Histograms of -$log_{10}$ p-values for lead SNPs for each “childhood adiposity” common factor in Models 1-4 (a-d) but which did not show genome-wide significant association ($P>5x10^{-8}$) with any individual contributing trait. The vertical red line indicates genome-wide significance. -log10(P)=-$log_{10}$ p-value for common “childhood adiposity” factor; AAM=age at menarche; SAC10=comparative body size age 10; MOBA7=childhood BMI age 7; MOBA8=childhood BMI age 8.*

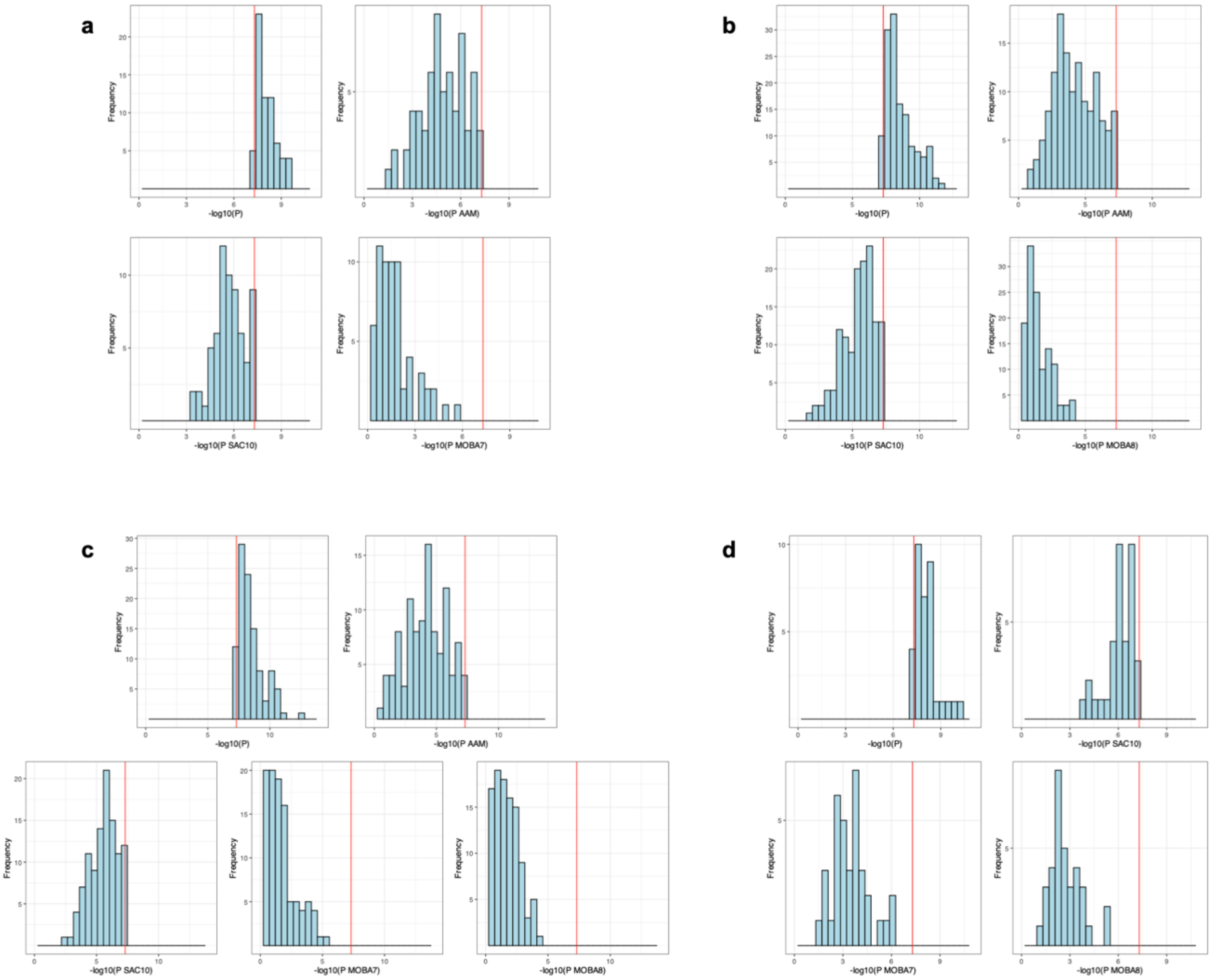

Exclusion of heterogenous GWAS signals: Some identified common factor signals may have disproportionately large effects on AAM and have relatively weak association with childhood BMI or SAC10. For each SNP, genomic SEM generates a Q index *p*-value ($p_Q$), indicating heterogeneity in its associations with the contributing individual traits. ~20% of signals across Models 1-3 (but none in Model 4) showed significant heterogeneity ($p_Q<5\times10^{-8}$); almost invariably such signals showed highly significant association with AAM but not with any individual childhood adiposity trait (**Figures 6**). The only exception was rs663129 (in Models 2 and 3), which showed genome-wide significant associations with SAC10, MOBA7 and MOBA8, but only nominal (and directionally inconsistent) association with AAM (**Figures 6b, 6c**). As rs663129 maps to *MC4R*, a known obesity gene, and showed strong associations with all adiposity phenotypes, we retained this signal. By contrast, other heterogenous SNPs mapped to well-known puberty genes such as *KISS1* and *TACR3* that are reported to be disrupted in monogenic disorders of puberty as well as several retinoic acid receptors involved in retinoic acid signalling (*RORB*, *RORA* and *RXRG*) and have previously been identified in GWAS for AAM. After excluding all heterogenous SNPs (except rs663129; n=117 SNPs):

- Model 1 (AAM, SAC10, MOBA7) had 400 signals
- Model 2 (AAM, SAC10, MOBA8) had 526 signals
- Model 3 (SAC10, MOBA7, MOBA8, AAM) had 440 signals
- Model 4 (SAC10, MOBA7, MOBA8) had 258 signals

***Figure 6* | *Scatterplots comparing common factor and individual trait associations (Z-scores) for the GWAS signals identified in Models 1-3 (a-c). Heterogenous signals ($p_Q$>5×10$^{-8}$) are highlighted in blue. The one retained heterogenous signal, rs663129, is labelled. AAM=age at menarche, SAC10=comparative body size age 10, MOBA7=childhood BMI age 7, MOBA8=childhood BMI age 8.***

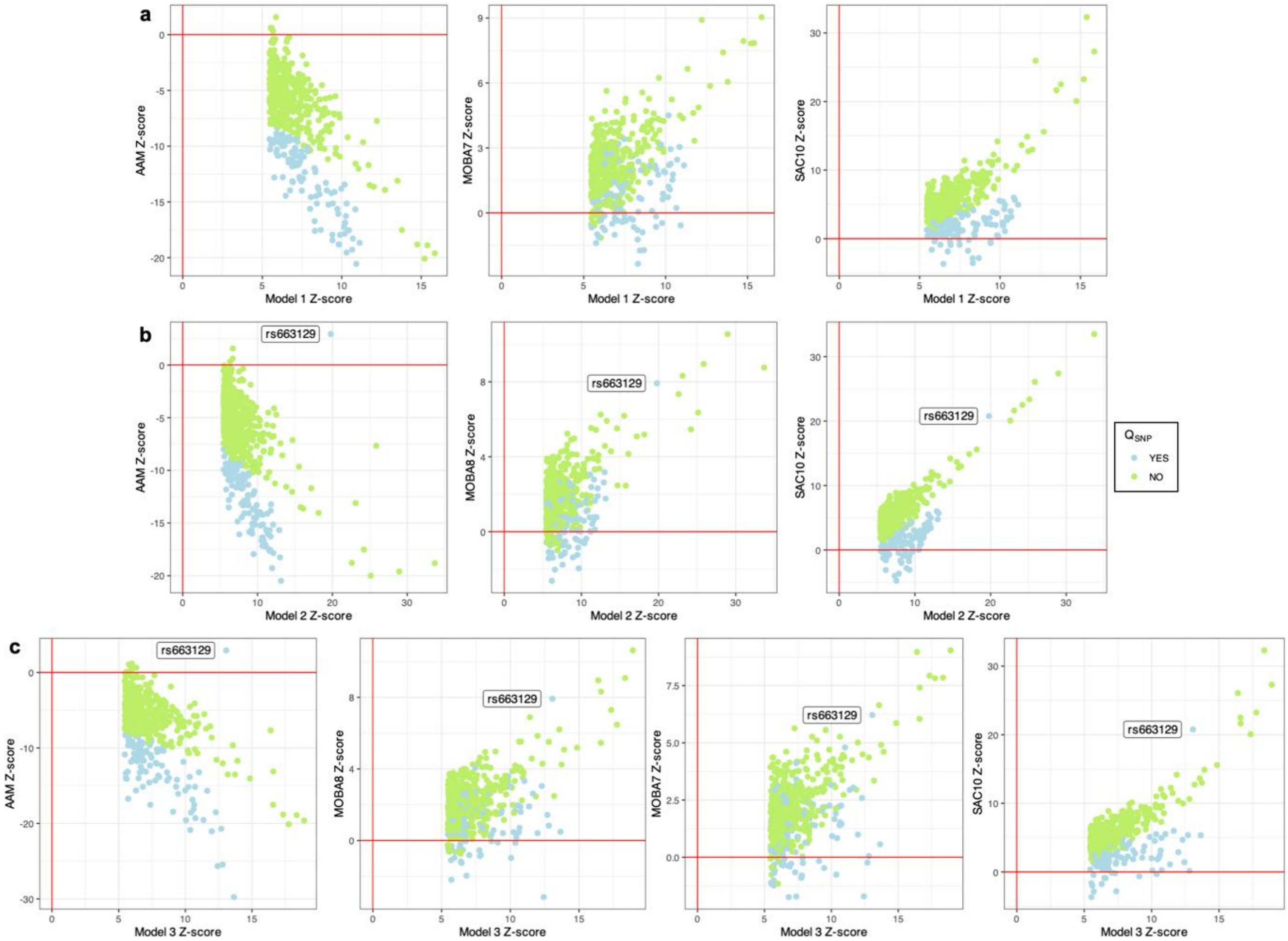

**Validity of GWAS signals for childhood adiposity common factors**

Genetic correlations with childhood BMI: The common factors from all four models showed high genetic correlations with childhood BMI from independent EGG data. Model 4 showed the highest genetic correlation with childhood BMI ($r_g$=0.97, 95% CI: 0.86-1.00) although its confidence intervals overlapped with the estimates from Models 1-3 ($r_g$=0.85-0.87).

Directional concordance with childhood BMI: After exclusion of heterogenous signals, GWAS signals for the common factors from all four models showed high directional concordance with effect estimates with childhood BMI. Signals from Model 4 showed the highest directional concordance (97%), and concordance was also high for signals from Models 1-3 (91-93%).

Concordance with adult BMI: Using data on adult BMI from the UK Biobank, we observed consistent moderate positive genetic correlations between all common factors and adult BMI ($r_g$=0.54-0.57). GWAS signals for the common factors from all four models showed consistently high directional concordance in effect estimates with adult BMI (93-96%).

In summary, leveraging AAM data increased the power to detect signals for a single "childhood adiposity" common factor (400-526 signals in Models 1-3 compared to 258 signals in Model 4). We selected Model 2 (AAM, SAC10, MOBA8) as it was the best-powered model, as well as performing well across all quality control and validation metrics.
